# DrugWAS: Leveraging drug-wide association studies to facilitate drug repurposing for COVID-19

**DOI:** 10.1101/2021.02.04.21251169

**Authors:** Cosmin A. Bejan, Katherine N. Cahill, Patrick J. Staso, Leena Choi, Josh F. Peterson, Elizabeth J. Phillips

## Abstract

**Importance:** There is an unprecedented need to rapidly identify safe and effective treatments for the novel coronavirus disease 2019 (COVID-19).

**Objective:** To systematically investigate if any of the available drugs in Electronic Health Record (EHR), including prescription drugs and dietary supplements, can be repurposed as potential treatment for COVID-19.

**Design, Setting, and Participants:** Based on a retrospective cohort analysis of EHR data, drug-wide association studies (DrugWAS) were performed on COVID-19 patients at Vanderbilt University Medical Center (VUMC). For each drug study, multivariable logistic regression with overlap weighting using propensity score was applied to estimate the effect of drug exposure on COVID-19 disease outcomes.

**Exposures:** Patient exposure to a drug during 1-year prior to the pandemic and COVID-19 diagnosis was chosen as exposure of interest. Natural language processing was employed to extract drug information from clinical notes, in addition to the prescription drug data available in structured format.

**Main Outcomes and Measures:** All-cause of death was selected as primary outcome. Hospitalization, admission to the intensive care unit (ICU), and need for mechanical ventilation were identified as secondary outcomes.

**Results:** The study included 7,768 COVID-19 patients, of which 509 (6.55%) were hospitalized, 82 (1.06%) were admitted to ICU, 64 (0.82%) received mechanical ventilation, and 90 (1.16%) died. Overall, 15 drugs were significantly associated with decreased COVID-19 severity. Previous exposure to either Streptococcus pneumoniae vaccines (adjusted odds ratio [OR], 0.38; 95% CI, 0.14-0.98), diphtheria toxoid vaccine (OR, 0.39; 95% CI, 0.15-0.98), and tetanus toxoid vaccine (OR, 0.39; 95% CI, 0.15-0.98) were significantly associated with a decreased risk of death (primary outcome). Secondary analyses identified several other significant associations showing lower risk for COVID-19 outcomes: 2 vaccines (acellular pertussis, Streptococcus pneumoniae), 3 dietary supplements (turmeric extract, flaxseed extract, omega-3 fatty acids), methylprednisolone acetate, pseudoephedrine, ethinyl estradiol, estradiol, ibuprofen, and fluticasone.

**Conclusions and Relevance:** This cohort study leveraged EHR data to identify a list of drugs that could be repurposed to improve COVID-19 outcomes. Further randomized clinical trials are needed to investigate the efficacy of the proposed drugs.

**Key Points:** *Question:* Can Electronic Health Records (EHRs) be used to search for drug candidates that could be repurposed to treat the coronavirus disease 2019 (COVID-19)?

*Findings:* Drug-wide association studies (DrugWAS) of COVID-19 severity outcomes were conducted on a cohort of 7,768 COVID-19 patients. The study found 15 drug ingredients that are significantly associated with a decreased risk of death and other severe COVID-19 outcomes.

*Meaning:* The list of drugs proposed by this study could provide additional insights into developing new candidates for COVID-19 treatment.

## 1. INTRODUCTION

Coronavirus disease 2019 (COVID-19), caused by severe acute respiratory syndrome coronavirus 2 (SARS-CoV-2), has triggered a pandemic infection leading to unprecedented excess mortality and adverse consequences to global economy.^1–3^ According to the World Health Organization (WHO), SARS-CoV-2 has spread to over 222 countries and territories resulting in >81 million infected individuals and >1.8 million confirmed deaths as of December 2020.^4^ While significant progress has been achieved to successfully develop and deploy safe and effective SARS-CoV-2 vaccines,^5–9^ intensive scientific efforts are currently underway to discover treatments that improve COVID-19 outcomes, particularly drug treatments that can be used early in a patient’s illness to prevent hospitalization or death. Recently, the antiviral drug remdesivir has been proven to reduce the recovery time of adult patients hospitalized with COVID-19.^10^ Another study indicated that use of dexamethasone reduces 28-day mortality of hospitalized COVID-19 patients receiving mechanical ventilation or high-flow oxygen.^11^ Monoclonal antibodies have been shown to reduce the viral load and improve clinical outcomes in outpatients with mild or moderate COVID-19.^12^ Furthermore, several other drugs including corticosteroids, antiviral therapies, immune-modulators, and anticoagulants are currently investigated as potential therapies for COVID-19.^13–17^ Despite recent advances, however, there is an urgent need for discovering safe and effective treatments that are able to prevent COVID-19 progression and long-term complications.^18^

Because a de novo treatment usually requires many years to reach the market, involves significant costs, and has a low rate of success, drug repurposing methodologies have emerged as an attractive strategy to accelerate the discovery of novel COVID-19 treatments.^19–21^ Leveraging real-world data from Electronic Health Record (EHR), we conducted a drug-wide association study (DrugWAS) to systematically investigate all recorded drug exposures, including prescription drugs and dietary supplements, as potential COVID-19 treatments. We hypothesized that drug exposures associated with a lower risk of death or severe COVID-19 outcomes could identify candidates for further therapeutic study.

## 2. METHODS

### Study Design

DrugWAS is a high-throughput method for independently investigating associations between drugs and disease outcomes. It relies on a retrospective cohort analysis of data stored in the Vanderbilt University Medical Center (VUMC) Research Derivative, a daily updated database of identified EHR data restructured for research. Specific data elements extracted from the Research Derivative include demographics data, laboratory tests, drugs, clinical outcomes, comorbidities, and clinical notes. The study was approved by the institutional review board at VUMC. It is presented by following the Strengthening the Reporting of Observational Studies in Epidemiology (STROBE) reporting guideline.^22^

### Study Population

The study included all patients who were tested at VUMC between March 9, 2020 and December 31, 2020 and were diagnosed with SARS-CoV-2 confirmed by polymerase chain reaction (PCR) assay (**Figure 1**). Being a large medical center in Middle Tennessee, and the sole provider of COVID-19 testing early in the pandemic, VUMC tested a substantial number of individuals who never had a care visit at the medical center prior to their test. As the baseline clinical and drug exposure data for these patients was sparse, and the risk of treatment misclassification was high, patients without an encounter in the EHR within a 1-year period prior to February 15, 2020 were excluded. The exclusion date corresponded to the date that COVID-19 pandemic was first detected in our geographical area. While no age, gender, race or ethnicity selection criteria were imposed, patients with missing demographic information (eg, unknown race) were excluded from the study. At VUMC, SARS-CoV-2 PCR testing was initially limited to symptomatic individuals and a selected category of patients who needed to be physically present in a VUMC facility (eg, pregnant women or patients scheduled for surgery) PCR testing was required before their visit. These patients, flagged as asymptomatic at the test time, were excluded from the study since they may increase the false positive rate of COVID-19 outcomes (eg, admission to hospital of an asymptomatic COVID-19 patient would be likely influenced by surgery rather than by the COVID-19 diagnosis).

**Figure 1.**
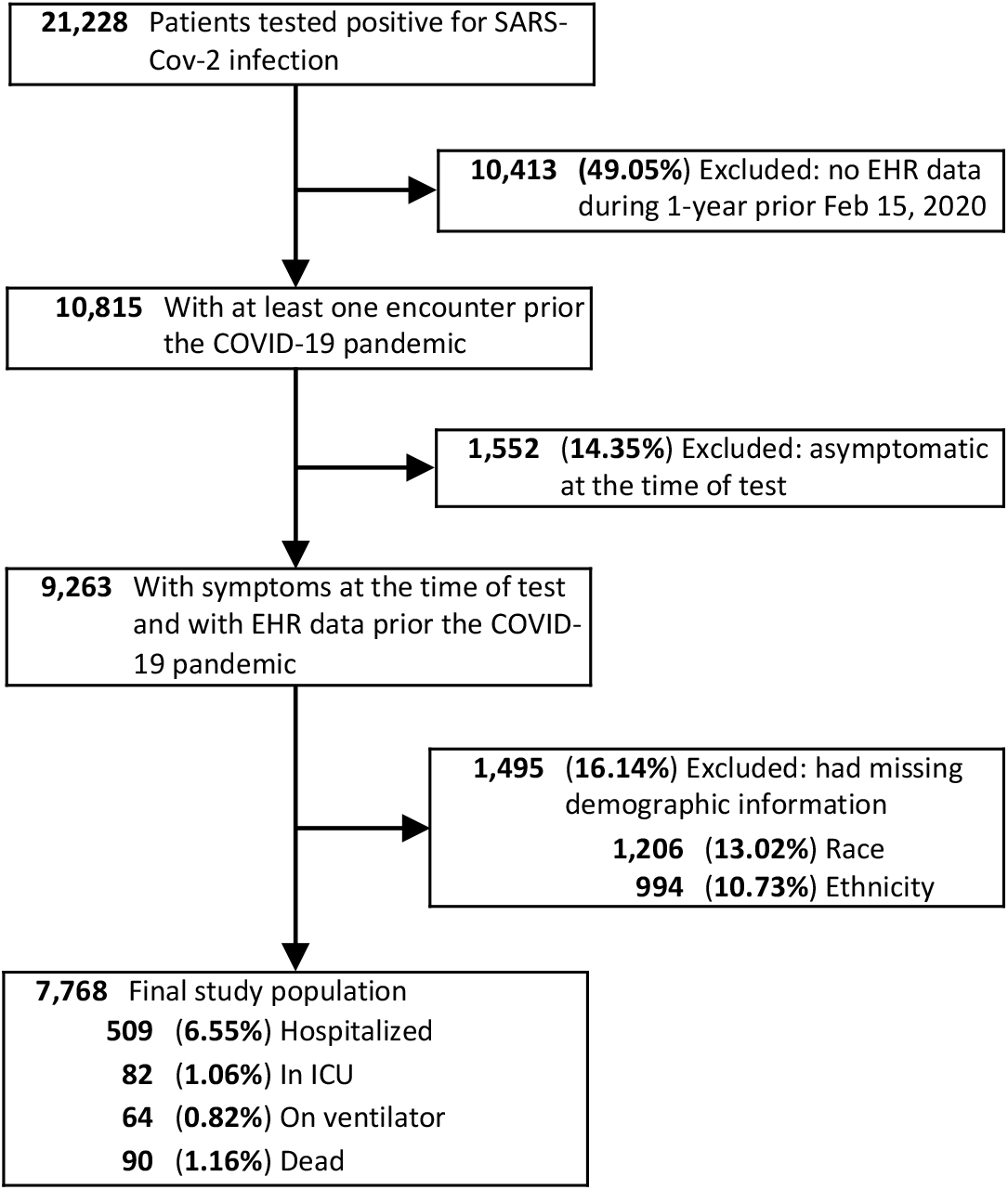
Selection of COVID-19 patients for the DrugWAS analysis.

### Exposure of Interest

Patient exposure to a drug between February 15, 2019 (approximated as 1-year prior to the pandemic arriving in our geographical area) and test time was selected as the exposure of interest for each drug-outcome association study in DrugWAS. All generic and brand drugs recorded in the EHR for the final study population during this interval were extracted from the drug table and normalized to drug ingredients using a previously developed drug normalization pipeline.^23^ Natural language processing (NLP) was also used to extract drug information from free-text notes. This process was particularly useful to identify exposure to over-the-counter drugs or to drugs prescribed by outside providers. For this, we employed MedXN-v1.0.3,^24^ a high-performance NLP drug extractor previously evaluated on Vanderbilt EHR,^25,26^ to parse >789,000 notes with dates between February 15, 2019 and PCR test time. There was no filter restriction by note type for NLP-based drug extraction; thus, notes such as problem lists, clinical communications, and outpatient Rx order summaries were also included in this process. Diagnostic drug ingredients, excipients, and other non-therapeutic agents (eg, placebo, inert ingredients) were excluded from the study. Drugs and supplements were assigned a therapeutic category (drug class) using the Lexicomp® database.

### Outcomes

Outcomes were extracted using EHR data from PCR test date until December 31, 2020. Based on the WHO guidelines on COVID-19 severity scale,^27^ they were classified as: 1) never hospitalized, 2) hospitalized with mild conditions and without intensive treatment (hospitalized-mild), 3) admitted to ICU, 4) on mechanical ventilation, and 5) dead. All-cause of death was selected as primary outcome. Two strategies were designed to combine hospitalized-mild, admitted to ICU, and on mechanical ventilation into multiple secondary outcomes: 1) cumulative severity, where a specific category is combined with more severe categories on the scale, and 2) exclusive severity, which includes only the patients from a specific category. For instance, the cumulative severity strategy for the ICU outcome also includes on ventilator and dead categories while the exclusive severity for the same outcome includes only the patients in ICU (**eFigure 1** in the Supplement). Of note, for the primary outcome, both strategies will generate the same severity group. All the severity groups corresponding to the primary and secondary outcomes were compared against non-hospitalized, alive COVID-19 patients.

### Covariates

Age, sex, race, and ethnicity were selected to account for the differences in patient characteristics. Additionally, the weighted Elixhauser comorbidity score^28,29^ was chosen to account for the severity of medical conditions since multiple comorbidities have been shown to be associated with COVID-19 outcomes.^30,31^ The covariate encoding this comorbidity score was computed by: 1) extracting the International Classification of Diseases, 9th/10th Revision, Clinical Modification (ICD-9/10-CM) billing codes from each patient record up to the PCR test time and determining their inclusion in any of the 31 Elixhauser comorbidity groups;^32^ 2) aggregating the severity scores derived from associations between the 31 comorbidity groups and the risk of in-hospital death;^29^ and 3) categorizing the aggregated severity scores into 4 ordinal categories: <0, 0, 1-4, and 5+.^33^

### Statistical Analysis

Patient characteristics were reported as means and standard deviations (SDs) for continuous variables and counts (percentages) for categorical variables.

For each drug studied, a propensity score method was used to adjust for differences between the patients exposed to the drug prior to being diagnosed with SARS-CoV-2 (exposed group) and those not exposed (unexposed group). The propensity score represents the probability of a patient being assigned to the exposed group conditional on the observed patient characteristics. In observational, nonrandomized studies, propensity score methods are used to balance the main patient characteristics across treatment groups, which is essential in reducing the bias in estimating treatment effects.^34–36^ In DrugWAS, the propensity score adjustment played a critical role in reducing the likelihood of confounding (especially confounding by indication) since patients exposed to drugs are likely to have comorbidities as treatment indications that may also affect the study outcomes.

This study applied the overlap weighing with a propensity score method. The method has been shown to achieve high performance under different configurations,^37,38^ and, recently, it has been successfully used in estimating the relationship between use of specific drugs and COVID-19 outcomes.^39–41^ Specifically, the propensity score for being exposed to a drug was estimated by a multivariable logistic regression model using age, sex, race, ethnicity, and weighted Elixhauser comorbidity score. Using the estimated propensity score, a weighted multivariable logistic regression was performed to estimate the effect of drug exposure on both primary and secondary outcomes, where each patient was weighted with the probability of the patient being assigned to the opposite exposure group. The estimates of adjusted odds ratio (OR) and 95% confidence intervals (CIs) were reported in the results. Associations corresponding to a specific outcome were performed for all drugs with at least 100 exposed patients of whom at least 5 had the outcome. All drugs with corresponding effect estimates indicating reduced severity risk (OR<1) were reported as potential candidates for COVID-19 treatment repurposing. No adjustment for multiple testing was performed. All statistical analyses were done in R, version 3.6.1.

## 3. RESULTS

### Patients

The study included 7,768 SARS-CoV-2 infected patients. The mean age was 42 and the majority of patients were females (61.3%), whites (84.2%), and non-Hispanic or Latino (96.5%). From this cohort, 509 (6.55%) were hospitalized and 90 (1.16%) died (**Figure 1**). Among those who died, 11 did not have an inpatient visit at VUMC after they were diagnosed with SARS-CoV-2 infection. While the weighted Elixhauser comorbidity score indicated a current state of health for most of the patients, 1,875 (24.1%) of them had severe comorbidity scores (**Table 1**). The hospitalized patients had a mean age of 60, and an increased percent of males (49.3%), blacks (24.8%), and severe comorbidity scores (58.7%).

**Table 1.** Patient characteristics.

| Characteristic | All patients |  | Hospitalized patients |  |
| --- | --- | --- | --- | --- |
|  | N | % | N | % |
| Total | 7,768 | 100 | 509 | 100 |
| Age, y* | 42 | 20 | 60 | 19 |
| Sex |  |  |  |  |
| Men | 3,003 | 38.7 | 251 | 49.3 |
| Women | 4,765 | 61.3 | 258 | 50.7 |
| Race |  |  |  |  |
| White | 6,543 | 84.2 | 369 | 72.5 |
| Black | 1,018 | 13.1 | 126 | 24.8 |
| Asian | 207 | 2.7 | 14 | 2.8 |
| Ethnicity |  |  |  |  |
| Not Hispanic or Latino | 7,497 | 96.5 | 488 | 95.9 |
| Hispanic or Latino | 271 | 3.5 | 21 | 4.1 |
| Weighted Elixhauser comorbidity score |  |  |  |  |
| <0 | 1,124 | 14.5 | 50 | 9.8 |
| 0 | 3,675 | 47.3 | 109 | 21.4 |
| 1-4 | 1,094 | 14.1 | 51 | 10 |
| 5+ | 1,875 | 24.1 | 299 | 58.7 |

\* Reported as age mean and standard deviation

### Primary Outcome

The analysis for the primary outcome consisted of 233 association studies between previous drug exposure and all-cause of death (**eTables 1** and **2** in the Supplement). Propensity score overlap-weighted logistic regression indicated that 49 drugs have lower death risk estimates (adjusted OR<1). Among these, previous exposures to Streptococcus pneumoniae serotype (1, 19A, 3, 5, 6A, 7F) capsular antigen diphtheria CRM197 protein conjugate vaccine (OR, 0.38; 95% CI, 0.14-0.98), Streptococcus pneumoniae serotype (14, 18C, 19F, 23F, 4, 6B, 9V) capsular antigen diphtheria CRM197 protein conjugate vaccine (OR, 0.38; 95% CI, 0.17-0.89), diphtheria toxoid vaccine (OR, 0.39; 95% CI, 0.15-0.98), and tetanus toxoid vaccine (OR, 0.39; 95% CI, 0.15-0.98) are associated with a significantly decreased risk of death. After overlap weighting with propensity score for the first (and second) Streptococcus pneumoniae vaccine the death rate was 2.1% (1.9%) in the exposed group compared with a death rate of 4.1% (3.6%) in the unexposed group. For both diphtheria and tetanus toxoid vaccines, the death rate was 0.7% vs 1.7% in the exposed and unexposed groups, respectively. **Figure 2** and **eTable 3** in the Supplement show additional details of the 49 drug studies for the primary outcome while **eTable 4** in the Supplement demonstrates that the overlap weighting achieved a good balance in main patients characteristics between the exposed and unexposed groups. A sensitivity analysis where the exposure of interest was extracted using only the information from the drug table (in structured format) found no significant associations for the primary outcome.

**Figure 2.**
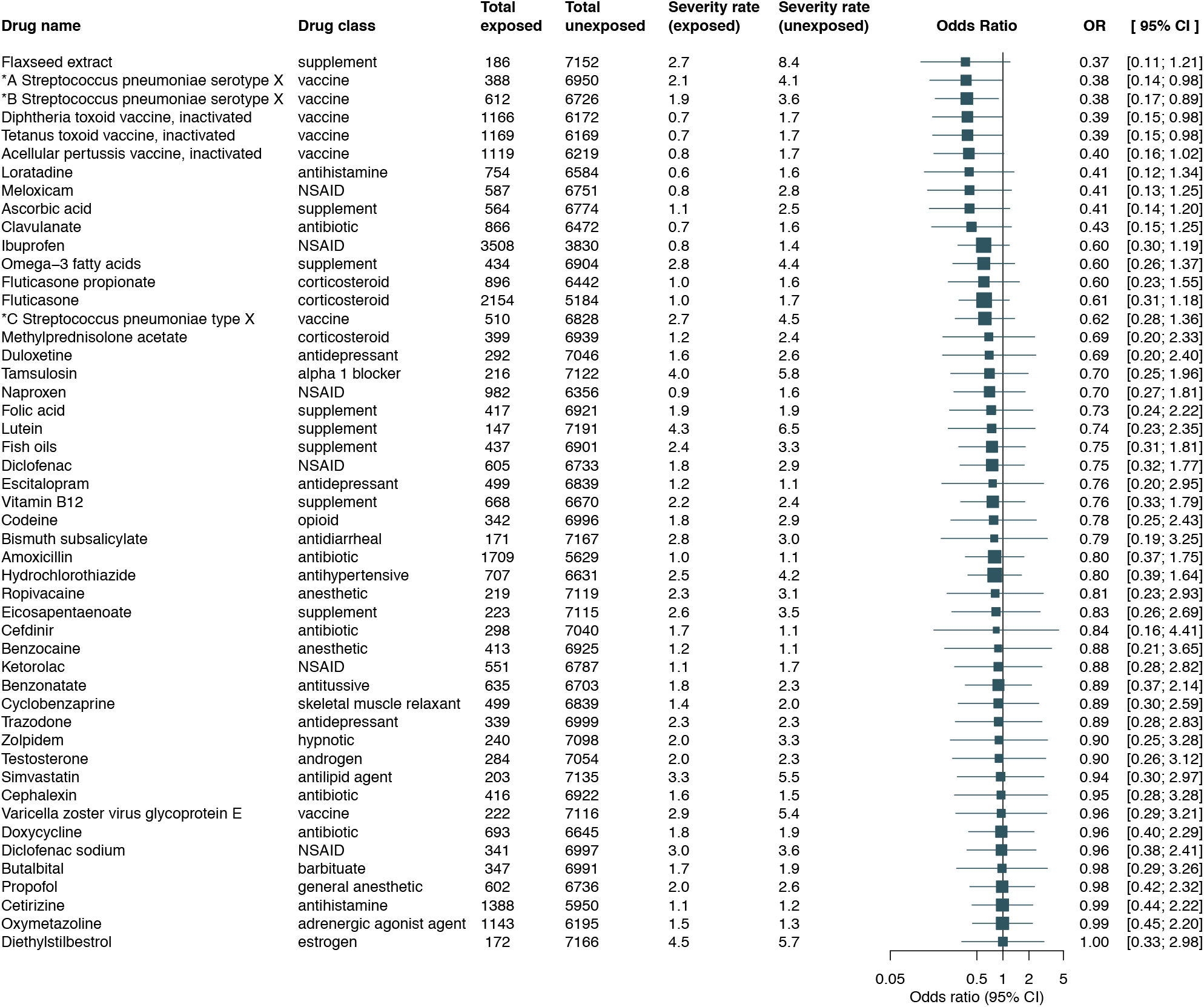
Association between drug exposure and all-cause of death. Adjusted odds ratios (ORs) and 95% confidence intervals (CIs) were estimated using propensity score overlap-weighted logistic regression while death rates were computed using overlap weighting with propensity score. Abbreviations and acronyms. ^*^A Streptococcus pneumoniae serotype X: Streptococcus pneumoniae serotype (1, 19A, 3, 5, 6A, 7F) capsular antigen diphtheria CRM197 protein conjugate vaccine; ^*^B Streptococcus pneumoniae serotype X: Streptococcus pneumoniae serotype (14, 18C, 19F, 23F, 4, 6B, 9V) capsular antigen diphtheria CRM197 protein conjugate vaccine; ^*^C Streptococcus pneumoniae type X: Streptococcus pneumoniae type (1, 10A, 11A, 12F, 14, 15B, 17F, 18C, 19A, 19F, 2, 20, 22F, 23F, 3, 33F, 4, 5, 6B, 7F, 8, 9N, 9V) capsular polysaccharide antigen; NSAID: nonsteroidal anti-inflammatory drug.

### Secondary Outcomes

The secondary outcome analyses lead to the discovery of additional drugs as potential treatments for COVID-19 (**eTables 5-10** in the Supplement). **Figure 3** summarizes the significant associations obtained across all COVID-19 outcomes. In addition to vaccines, pervious exposure to flaxseed extract, methylprednisolone acetate, pseudoephedrine, omega-3 fatty acids, turmeric extract, ibuprofen, and fluticasone showed significant lower risks for hospitalized-mild (cumulative and exclusive severity). Furthermore, ethinyl estradiol and estradiol demonstrated protective effect for hospitalized-mild (cumulative severity).

**Figure 3.**
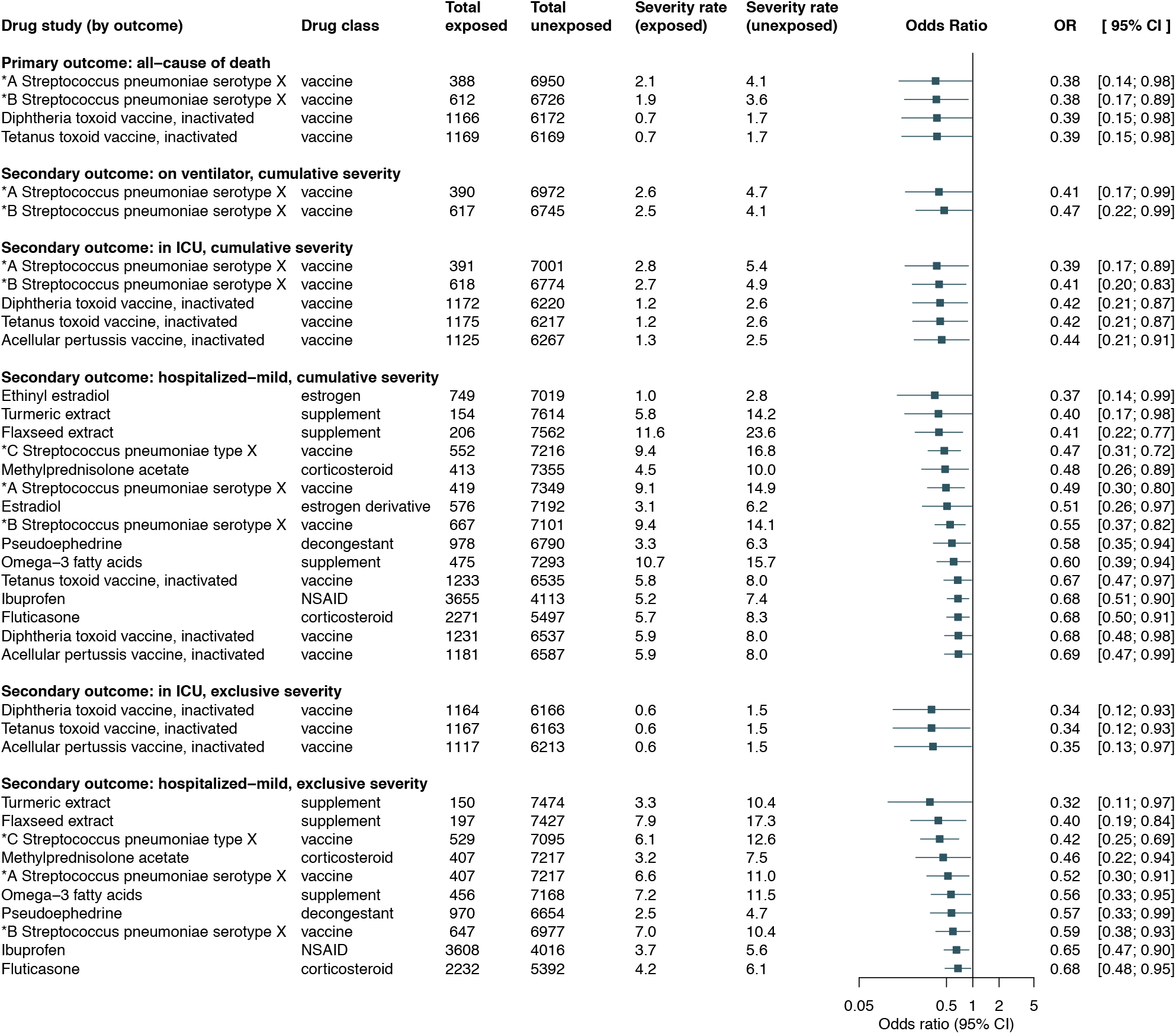
Significant associations grouped by outcome. Abbreviations and acronyms. ^*^A Streptococcus pneumoniae serotype X: Streptococcus pneumoniae serotype (1, 19A, 3, 5, 6A, 7F) capsular antigen diphtheria CRM197 protein conjugate vaccine; ^*^B Streptococcus pneumoniae serotype X: Streptococcus pneumoniae serotype (14, 18C, 19F, 23F, 4, 6B, 9V) capsular antigen diphtheria CRM197 protein conjugate vaccine; ^*^C Streptococcus pneumoniae type X: Streptococcus pneumoniae type (1, 10A, 11A, 12F, 14, 15B, 17F, 18C, 19A, 19F, 2, 20, 22F, 23F, 3, 33F, 4, 5, 6B, 7F, 8, 9N, 9V) capsular polysaccharide antigen; NSAID: nonsteroidal anti-inflammatory drug.

### Risk Trend Analysis

A visual analysis showing the risk trends over the selected outcomes for all 49 drugs from the primary analysis is depicted in **Figure 4**. Spline regression was also used to visualize non-linearity of risk trends across COVID-19 outcomes ordered by increased severity (**eFigures 2** and **3** in the Supplement). Drugs including fluticasone, naproxen, flaxseed extract, and omega-3 fatty acids show a constant low risk across the entire COVID-19 severity scale whereas cephalexin and cyclobenzaprine indicate a protective effect only for the primary outcome.

**Figure 4.**
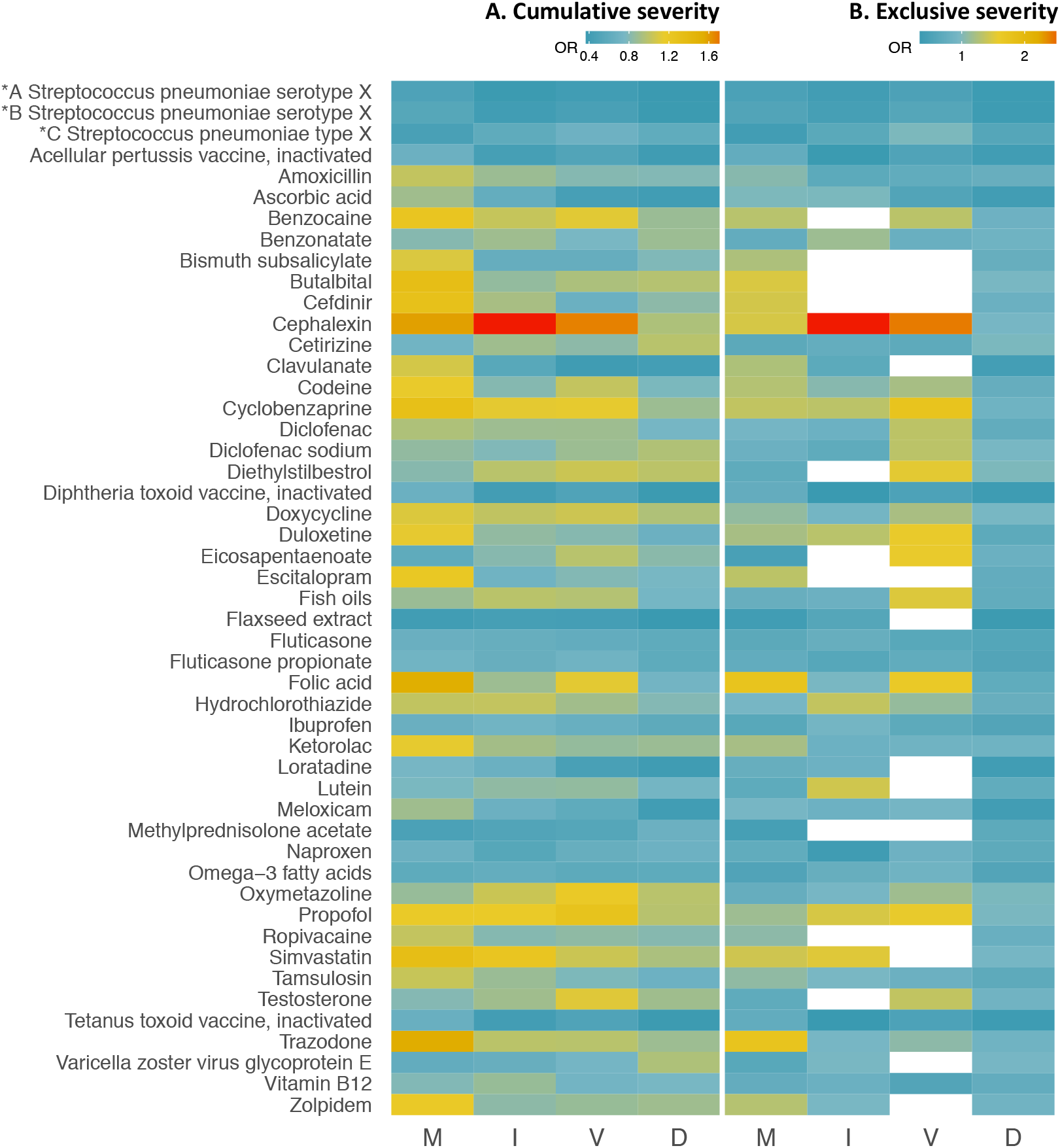
Heatmap of 49 drug associations across the COVID-19 severity scale: hospitalized-mild (M), ICU admission (I), on ventilation (V), and death (D). In Figures 4A and 4B the secondary outcomes were extracted following the cumulative severity and exclusive severity strategies, respectively. In Figure 4B, drug studies that did not meet the inclusion criteria are shown in white. Abbreviations: ^*^A Streptococcus pneumoniae serotype X: Streptococcus pneumoniae serotype (1, 19A, 3, 5, 6A, 7F) capsular antigen diphtheria CRM197 protein conjugate vaccine; ^*^B Streptococcus pneumoniae serotype X: Streptococcus pneumoniae serotype (14, 18C, 19F, 23F, 4, 6B, 9V) capsular antigen diphtheria CRM197 protein conjugate vaccine; ^*^C Streptococcus pneumoniae type X: Streptococcus pneumoniae type (1, 10A, 11A, 12F, 14, 15B, 17F, 18C, 19A, 19F, 2, 20, 22F, 23F, 3, 33F, 4, 5, 6B, 7F, 8, 9N, 9V) capsular polysaccharide antigen

## 4. DISCUSSION

We presented a high-throughput method to systematically investigate associations between drug exposures and COVID-19 outcomes. Our study found 15 drug ingredients that are significantly associated with a decreased risk of death and other severe COVID-19 outcomes. Moreover, 103 other drug ingredients indicated a protective effect for COVID-19 outcomes. It is our hope that this proposed list of drug ingredients provides additional insights into developing efficient COVID-19 treatments and would serve as a starting point for future prospective studies. Since their short- and long-term adverse events have been already studied, the efficacy of these drug ingredients against COVID-19 could be investigated rapidly in clinical trials.^42^

For each drug study, overlap weighting with propensity score was implemented to adjust for confounding when comparing the drug exposed and unexposed groups. Another significant contribution of our study is the use of NLP to better capture drug exposure information from clinical notes. Notably, this process had a critical role in enabling the study of over-the-counter drugs including dietary supplements as potential COVID-19 treatments.

Some of the significant drug ingredients found in this study replicate previous findings of potential dug repurposing candidates for COVID-19. Two network-based bioinformatics approaches found fluticasone as a possible efficient treatment for COVID-19^20,43^ while an in vitro study indicated that fluticasone does not suppress SARS-CoV-2 replication.^44^ Methylprednisolone has been previously shown to be associated with a decreased risk of death.^30^ It is currently recommended as a treatment for COVID-19 patients with severe and critical outcomes when dexamethasone is unavailable.^45^ In a network-based bioinformatics analysis pseudoephedrine was ranked as the best treatment candidate against COVID-19.^46^ Concerns regarding the use of ibuprofen causing potential harm to COVID-19 patients has initially received significant attention.^47–49^ More recent studies, however, found no significant evidence to suggest that ibuprofen is associated with severe COVID-19 outcomes.^50,51^ Further, an observational study using EHR data from 6 hospitals indicated that exposure to ibuprofen is associated with a lower risk of hospitalization due to COVID-19 (OR, 0.73; 95% CI, 0.64-0.84).^52^ A retrospective study using EHR data showed a decreased fatality rate for women 50+ years old receiving estradiol therapy (OR, 0.33; 95% CI, 0.18-0.62).^53^ Multiple clinical trials are currently underway to test the efficacy and safety of using methylprednisolone (NCT04263402, NCT04559113, NCT04636671), ibuprofen (NCT04334629, NCT04382768, NCT04383899), and estradiol (NCT04539626) against COVID-19.

An important finding by our study is that recent exposure to various types of Streptococcus pneumoniae vaccines, diphtheria toxoid vaccine, tetanus toxoid vaccine, and acellular pertussis vaccine is associated with a decreased risk of death and other severe COVID-19 outcomes. This may be explained by the ability of these vaccines to stimulate the immune system and provide immunologic protection against SARS-CoV-2.^54^ Pneumonia and influenza vaccination was also suggested to prevent COVID-19 exacerbation due to co-infection with other viruses.^55^ To our knowledge, dietary supplements such as flaxseed extract, omega-3 fatty acids, and turmeric extract have not been previously shown to be associated with a reduced COVID-19 severity risk. However, due to their anti-inflammatory properties, they have been proposed as alternative treatments to improve the clinical outcomes of COVID-19 patients.^56–61^ Yet, the dietary supplement results should be interpreted with caution since the corresponding ingredients are not evaluated by the United States Food and Drug Administration for safety and effectiveness and are not intended to diagnose, treat, cure, or prevent any disease.

### Limitations

Although overlap weighting with propensity score was applied to balance out the main patient characteristics between the drug exposed and unexposed groups, there may be unmeasured confounding factors that were not included in the propensity score model. Thus, while the bias may be reduced, confounding by indication is still possible due to unmeasured confounding factors. This is a major limitation, generally applicable to observational studies that lack randomization for drug exposure assignment.^40,62^

Treatment misclassification was addressed by including only patients with at least one encounter in the EHR such that drug exposure information could be extracted for each SARS-CoV-2 positive patient during at least one year prior to diagnosis. However, EHR phenotyping pose many challenges which could lead to inaccurately extracting the treatment status of the patient.^63^ For example, drug exposure extraction from clinical text relies on accurate identification of text expressions that are negated (eg, “*he could not be on Coumadin because of history of GI bleed*”) or hypothetical (eg, “*Zofran 4 mg PO once a day as needed for nausea*”).^23,64^ This is another reason for interpreting the results of dietary supplements with caution since they are primarily extracted from clinical notes. Furthermore, assuming drug exposure information is accurately extracted for a patient, exposure of the drug at and after diagnosis time is not guaranteed. However, vaccine data in the EHR is not subject to this limitation.

Misclassification of COVID-19 outcomes was addressed by excluding patients that were asymptomatic at test time. These patients could introduce bias in the study since, for instance, they may be admitted to hospital for reasons other than COVID-19. Additionally, despite including only patients with VUMC as their “medical home”, there could be a small number of patients among those classified as non-hospitalized who were in fact admitted to another hospital.

Finally, generalizability has yet to be proven on larger cohorts with a more heterogeneous study population. In this single-site study, patients are predominantly white, non-Hispanic or Latino and relatively young (mean age of 42). Larger cohorts would also enable conducting subgroup analyses for a specific race, sex, age category, set of comorbidities, drug dose, drug route, drug exposure intervals, or combination therapy.

## 5. CONCLUSIONS

Leveraging EHR data, DrugWAS of COVID-19 severity outcomes enables the discovery of drug ingredients that could be repurposed as potential treatments for COVID-19. In addition to the prescription drugs available in structured format, extracting drug information from clinical notes using NLP facilitates the study of over-the-counter drugs on improving the recovery of COVID-19 patients. The efficacy of the identified drug ingredients needs to be evaluated in prospective clinical trials using larger and more heterogenous study populations.

## Data Availability

Data contain protected health information and are not publicly available. The summary statistics extracted from the EHR data used in this study are provided in the manuscript and supplementary material.

## Author Contributions

Dr Bejan had full access to all the data in the study and takes responsibility for the integrity of the data and the accuracy of the data analysis. Study concept and design: Bejan, Cahill, Phillips. Acquisition, analysis, or interpretation of data: All authors. Drafting of the manuscript: Bejan. Critical revision of the manuscript for important intellectual content: Bejan, Cahill, Choi, Peterson, Phillips. Statistical analysis: Bejan. Administrative, technical, or material support: Bejan, Cahill, Staso. Study supervision: Phillips.

