## Supplementary Material for "DrugWAS: Leveraging drug-wide association studies to facilitate drug repurposing for COVID-19"

### Supplemental Figures

**eFigure 1** Strategies for selecting severity groups for COVID-19 outcomes

**eFigure 2** Spline regression analysis showing the OR trends of 49 drug associations across the COVID-19 severity scale (cumulative severity strategy)

**eFigure 3** Spline regression analysis showing the OR trends of 49 drug associations across the COVID-19 severity scale (exclusive severity strategy)

### Supplemental Tables

**eTable 1** The number of total, potential, and significant associations conducted for primary and secondary outcomes

**eTable 2** Patient counts for each severity group included in the study

**eTable 3** Association between drug exposure and all-cause of death

**eTable 4** Main patient characteristics by overlap weighting with propensity scores for all drug studies in the primary analysis

**eTable 5** Association between drug exposure and on ventilator (cumulative severity)

**eTable 6** Association between drug exposure and in ICU (cumulative severity)

**eTable 7** Association between drug exposure and hospitalized-mild (cumulative severity)

**eTable 8** Association between drug exposure and on ventilator (exclusive severity)

**eTable 9** Association between drug exposure and in ICU (exclusive severity)

**eTable 10** Association between drug exposure and hospitalized-mild (exclusive severity)

### Supplemental Figures

**eFigure 1** Strategies for selecting severity groups for COVID-19 outcomes. **A.** Cumulative severity strategy for the in ICU outcome includes in the severity group all the patients admitted to ICU or with a more severe outcome (ie, on ventilator or dead). **B.** Exclusive severity for the in ICU outcomes includes in the severity group only the patients admitted to ICU and exclude all the patients with any other outcome.

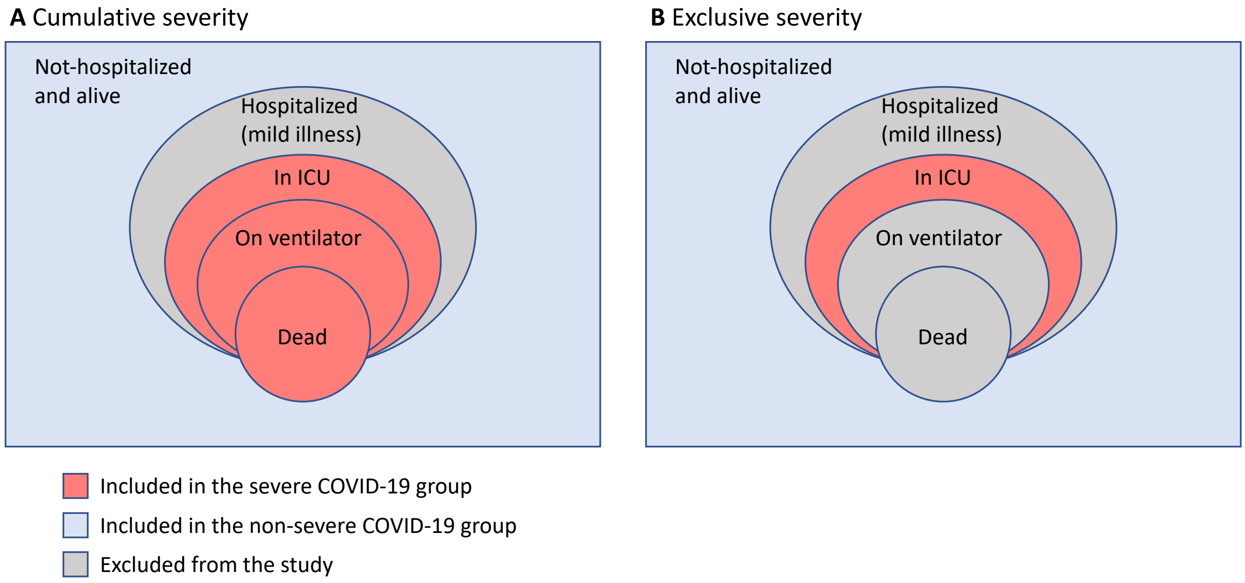

**eFigure 2** Spline regression analysis showing the OR trends of 49 drug associations across the COVID-19 severity scale: hospitalized-mild (M), ICU admission (I), on ventilation (V), and death (D). The secondary outcomes were extracted following the cumulative severity strategy.

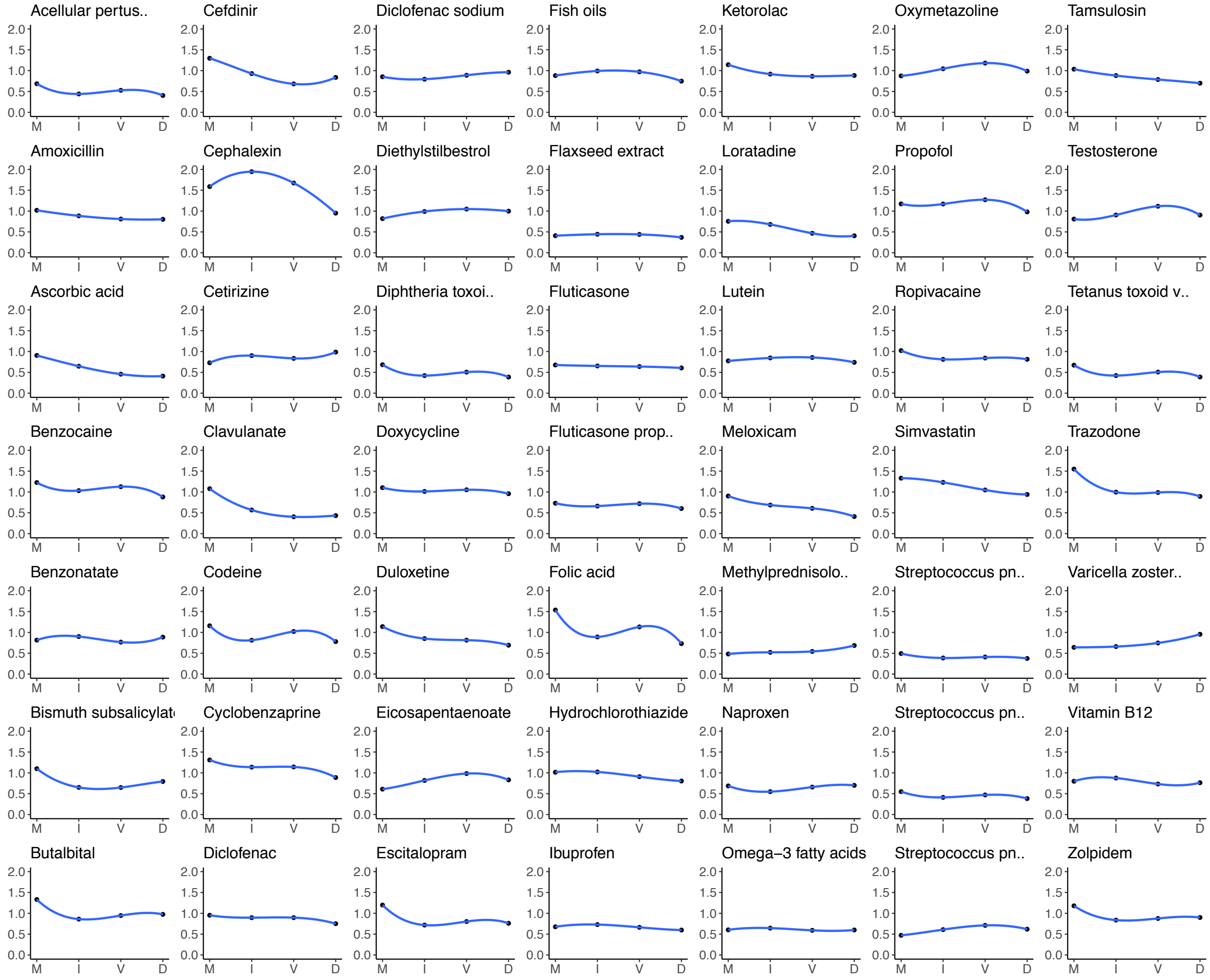

**eFigure 3** Spline regression analysis showing the OR trends of 49 drug associations across the COVID-19 severity scale: hospitalized-mild (M), ICU admission (I), on ventilation (V), and death (D). The secondary outcomes were extracted following the exclusive severity strategy.

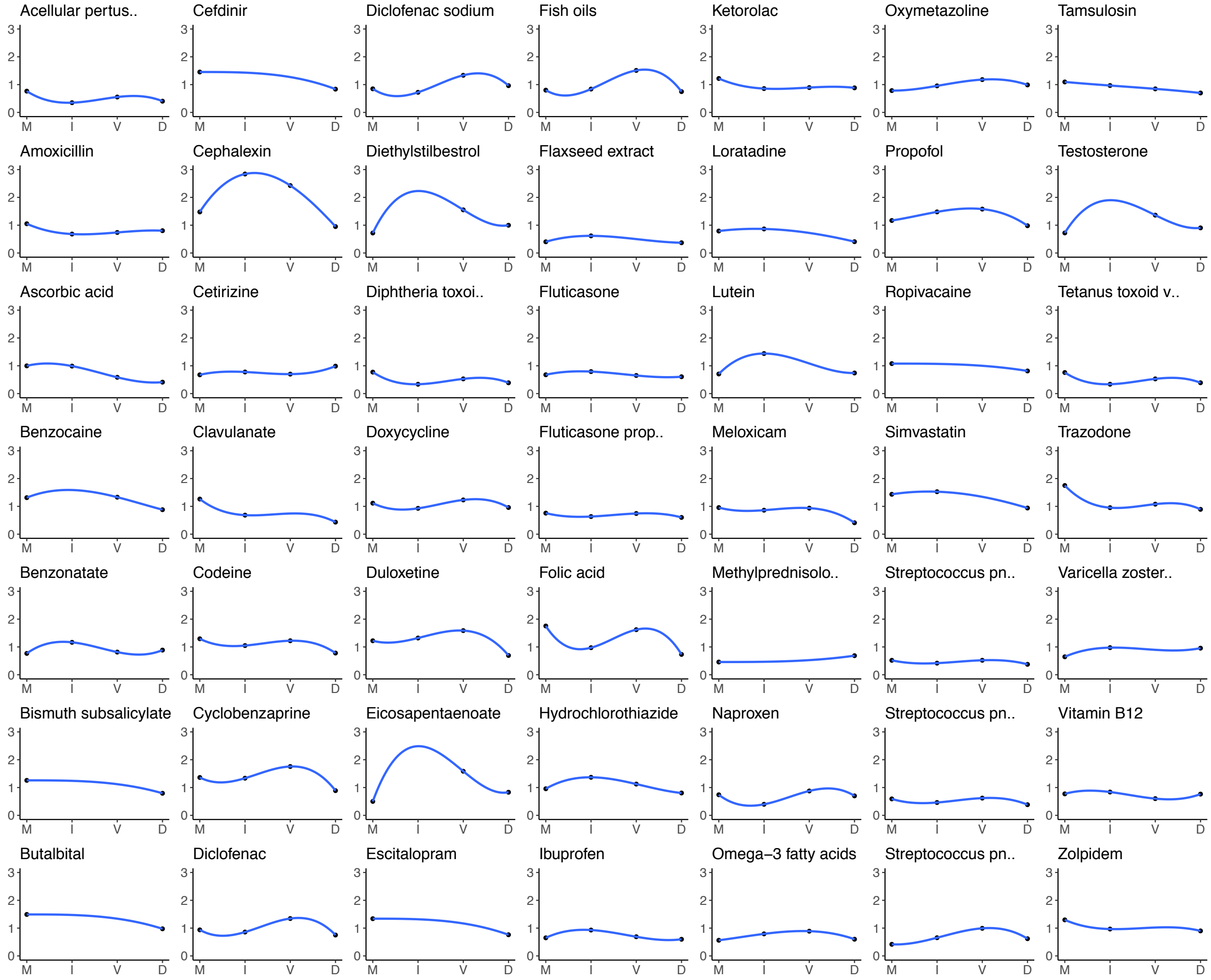

### Supplemental Tables

**eTable 1** The number of total, potential, and significant associations conducted for primary and secondary outcomes.

| | Cumulative severity | | | Exclusive severity | | |
|---|---|---|---|---|---|---|
| Study | Significant | Potential | Total | Significant | Potential | Total |
| Dead | 4 | 49 | 233 | 4 | 49 | 233 |
| On ventilator | 2 | 48 | 243 | 0 | 24 | 198 |
| In ICU | 5 | 61 | 261 | 3 | 45 | 202 |
| Hospitalized-mild | 15 | 77 | 346 | 10 | 73 | 331 |

**eTable 2** Patient counts for each severity group included in the study. Notably, the hospitalized-mild cohort extracted using the cumulative severity strategy includes all hospitalized patients (N=509) and the patients who died and did not have an inpatient visit at VUMC after they were diagnosed with SARS-CoV-2 infection (N=11).

| Severity group | Cumulative severity | Exclusive severity |
|---|---|---|
| Dead | 90 | 90 |
| On ventilator | 114 | 64 |
| In ICU | 144 | 82 |
| Hospitalized-mild | 520 | 376 |
| Not hospitalized, alive | 7,248 | 7,248 |

**eTable 3** Association between drug exposure and all-cause of death.

**TCe/u**, total count of drug exposed/unexposed patients; **SRe/u**, severity rate of drug exposed/unexposed patients computed after overlap weighting with propensity score; **OR**, odds ratio; **CI**, confidence interval. The association results are arranged in the ascending order of OR values. The statistically significant results at the 0.05 level are emphasized in bold.

| Drug name | Drug class | TCe | TCu | SRe | SRu | OR | 95% CI |
|---|---|---|---|---|---|---|---|
| Flaxseed extract | supplement | 186 | 7,152 | 2.7 | 8.4 | 0.37 | (0.11 - 1.21) |
| ***A Streptococcus pneumoniae serotype X** | **vaccine** | **388** | **6,950** | **2.1** | **4.1** | **0.38** | **(0.14 - 0.98)** |
| ***B Streptococcus pneumoniae serotype X** | **vaccine** | **612** | **6,726** | **1.9** | **3.6** | **0.38** | **(0.17 - 0.89)** |
| **Diphtheria toxoid vaccine, inactivated** | **vaccine** | **1,166** | **6,172** | **0.7** | **1.7** | **0.39** | **(0.15 - 0.98)** |
| **Tetanus toxoid vaccine, inactivated** | **vaccine** | **1,169** | **6,169** | **0.7** | **1.7** | **0.39** | **(0.15 - 0.98)** |
| Acellular pertussis vaccine, inactivated | vaccine | 1,119 | 6,219 | 0.8 | 1.7 | 0.4 | (0.16 - 1.02) |
| Loratadine | antihistamine | 754 | 6,584 | 0.6 | 1.6 | 0.41 | (0.12 - 1.34) |
| Meloxicam | NSAID | 587 | 6,751 | 0.8 | 2.8 | 0.41 | (0.13 - 1.25) |
| Ascorbic acid | supplement | 564 | 6,774 | 1.1 | 2.5 | 0.41 | (0.14 - 1.20) |
| Clavulanate | antibiotic | 866 | 6,472 | 0.7 | 1.6 | 0.43 | (0.15 - 1.25) |
| Ibuprofen | NSAID | 3,508 | 3,830 | 0.8 | 1.4 | 0.6 | (0.30 - 1.19) |
| Omega-3 fatty acids | supplement | 434 | 6,904 | 2.8 | 4.4 | 0.6 | (0.26 - 1.37) |
| Fluticasone propionate | corticosteroid | 896 | 6,442 | 1 | 1.6 | 0.6 | (0.23 - 1.55) |
| Fluticasone | corticosteroid | 2,154 | 5,184 | 1 | 1.7 | 0.61 | (0.31 - 1.18) |
| *C Streptococcus pneumoniae type X | vaccine | 510 | 6,828 | 2.7 | 4.5 | 0.62 | (0.28 - 1.36) |
| Methylprednisolone acetate | corticosteroid | 399 | 6,939 | 1.2 | 2.4 | 0.69 | (0.20 - 2.33) |
| Duloxetine | antidepressant | 292 | 7,046 | 1.6 | 2.6 | 0.69 | (0.20 - 2.40) |
| Tamsulosin | alpha 1 blocker | 216 | 7,122 | 4 | 5.8 | 0.7 | (0.25 - 1.96) |
| Naproxen | NSAID | 982 | 6,356 | 0.9 | 1.6 | 0.7 | (0.27 - 1.81) |
| Folic acid | supplement | 417 | 6,921 | 1.9 | 1.9 | 0.73 | (0.24 - 2.22) |
| Lutein | supplement | 147 | 7,191 | 4.3 | 6.5 | 0.74 | (0.23 - 2.35) |
| Fish oils | supplement | 437 | 6,901 | 2.4 | 3.3 | 0.75 | (0.31 - 1.81) |
| Diclofenac | NSAID | 605 | 6,733 | 1.8 | 2.9 | 0.75 | (0.32 - 1.77) |
| Escitalopram | antidepressant | 499 | 6,839 | 1.2 | 1.1 | 0.76 | (0.20 - 2.95) |
| Vitamin B12 | supplement | 668 | 6,670 | 2.2 | 2.4 | 0.76 | (0.33 - 1.79) |
| Codeine | opioid | 342 | 6,996 | 1.8 | 2.9 | 0.78 | (0.25 - 2.43) |
| Bismuth subsalicylate | antidiarrheal | 171 | 7,167 | 2.8 | 3 | 0.79 | (0.19 - 3.25) |
| Amoxicillin | antibiotic | 1,709 | 5,629 | 1 | 1.1 | 0.8 | (0.37 - 1.75) |
| Hydrochlorothiazide | antihypertensive | 707 | 6,631 | 2.5 | 4.2 | 0.8 | (0.39 - 1.64) |
| Ropivacaine | anesthetic | 219 | 7,119 | 2.3 | 3.1 | 0.81 | (0.23 - 2.93) |
| Eicosapentaenoate | supplement | 223 | 7,115 | 2.6 | 3.5 | 0.83 | (0.26 - 2.69) |
| Cefdinir | antibiotic | 298 | 7,040 | 1.7 | 1.1 | 0.84 | (0.16 - 4.41) |
| Benzocaine | anesthetic | 413 | 6,925 | 1.2 | 1.1 | 0.88 | (0.21 - 3.65) |
| Ketorolac | NSAID | 551 | 6,787 | 1.1 | 1.7 | 0.88 | (0.28 - 2.82) |

| | | | | | | | |
|---|---|---|---|---|---|---|---|
| Benzonatate | antitussive | 635 | 6,703 | 1.8 | 2.3 | 0.89 | (0.37 - 2.14) |
| Cyclobenzaprine | skeletal muscle relaxant | 499 | 6,839 | 1.4 | 2 | 0.89 | (0.30 - 2.59) |
| Trazodone | antidepressant | 339 | 6,999 | 2.3 | 2.3 | 0.89 | (0.28 - 2.83) |
| Zolpidem | hypnotic | 240 | 7,098 | 2 | 3.3 | 0.9 | (0.25 - 3.28) |
| Testosterone | androgen | 284 | 7,054 | 2 | 2.3 | 0.9 | (0.26 - 3.12) |
| Simvastatin | antilipid agent | 203 | 7,135 | 3.3 | 5.5 | 0.94 | (0.30 - 2.97) |
| Cephalexin | antibiotic | 416 | 6,922 | 1.6 | 1.5 | 0.95 | (0.28 - 3.28) |
| Varicella zoster virus glycoprotein E | vaccine | 222 | 7,116 | 2.9 | 5.4 | 0.96 | (0.29 - 3.21) |
| Doxycycline | antibiotic | 693 | 6,645 | 1.8 | 1.9 | 0.96 | (0.40 - 2.29) |
| Diclofenac sodium | NSAID | 341 | 6,997 | 3 | 3.6 | 0.96 | (0.38 - 2.41) |
| Butalbital | barbituate | 347 | 6,991 | 1.7 | 1.9 | 0.98 | (0.29 - 3.26) |
| Propofol | general anesthetic | 602 | 6,736 | 2 | 2.6 | 0.98 | (0.42 - 2.32) |
| Cetirizine | antihistamine | 1,388 | 5,950 | 1.1 | 1.2 | 0.99 | (0.44 - 2.22) |
| Oxymetazoline | adrenergic agonist agent | 1,143 | 6,195 | 1.5 | 1.3 | 0.99 | (0.45 - 2.20) |
| Diethylstilbestrol | estrogen | 172 | 7,166 | 4.5 | 5.7 | 1 | (0.33 - 2.98) |

Other abbreviations and acronyms. *A Streptococcus pneumoniae serotype X: Streptococcus pneumoniae serotype (1, 19A, 3, 5, 6A, 7F) capsular antigen diphtheria CRM197 protein conjugate vaccine; *B Streptococcus pneumoniae serotype X: Streptococcus pneumoniae serotype (14, 18C, 19F, 23F, 4, 6B, 9V) capsular antigen diphtheria CRM197 protein conjugate vaccine; *C Streptococcus pneumoniae type X: Streptococcus pneumoniae type (1, 10A, 11A, 12F, 14, 15B, 17F, 18C, 19A, 19F, 2, 20, 22F, 23F, 3, 33F, 4, 5, 6B, 7F, 8, 9N, 9V) capsular polysaccharide antigen; NSAID: nonsteroidal anti-inflammatory drug.

**eTable 4** Main patient characteristics by overlap weighting with propensity scores for all drug studies in the primary analysis.

| Drug name | Drug exposed | Patient count | Male, % | White, % | Black, % | Not Hispanic or Latino, % |
|---|---|---|---|---|---|---|
| Flaxseed extract | yes | 186 | 43.6 | 92.1 | 7.9 | 97.5 |
| | no | 7,152 | 43.6 | 92.1 | 7.9 | 97.5 |
| *A Streptococcus pneumoniae serotype X | yes | 388 | 42.8 | 83.8 | 12 | 94.4 |
| | no | 6,950 | 42.8 | 83.8 | 12 | 94.4 |
| *B Streptococcus pneumoniae serotype X | yes | 612 | 44.8 | 81.7 | 14.5 | 95.3 |
| | no | 6,726 | 44.8 | 81.7 | 14.5 | 95.3 |
| Diphtheria toxoid vaccine, inactivated | yes | 1,166 | 38.1 | 82.2 | 14.1 | 95.6 |
| | no | 6,172 | 38.1 | 82.2 | 14.1 | 95.6 |
| Tetanus toxoid vaccine, inactivated | yes | 1,169 | 38.2 | 82.2 | 14.1 | 95.6 |
| | no | 6,169 | 38.2 | 82.2 | 14.1 | 95.6 |
| Acellular pertussis vaccine, inactivated | yes | 1,119 | 37.8 | 81.8 | 14.5 | 95.7 |
| | no | 6,219 | 37.8 | 81.8 | 14.5 | 95.7 |
| Loratadine | yes | 754 | 35.3 | 85.5 | 11.5 | 95.9 |
| | no | 6,584 | 35.3 | 85.5 | 11.5 | 95.9 |
| Meloxicam | yes | 587 | 33.3 | 86 | 12.4 | 97.2 |
| | no | 6,751 | 33.3 | 86 | 12.4 | 97.2 |
| Ascorbic acid | yes | 564 | 32.4 | 87.4 | 9.9 | 97.1 |
| | no | 6,774 | 32.4 | 87.4 | 9.9 | 97.1 |
| Clavulanate | yes | 866 | 33.7 | 86.3 | 12.1 | 96.9 |
| | no | 6,472 | 33.7 | 86.3 | 12.1 | 96.9 |
| Ibuprofen | yes | 3,508 | 37.6 | 85.1 | 12.3 | 96.6 |
| | no | 3,830 | 37.6 | 85.1 | 12.3 | 96.6 |
| Omega-3 fatty acids | yes | 434 | 43.1 | 91 | 6.7 | 97.3 |
| | no | 6,904 | 43.1 | 91 | 6.7 | 97.3 |
| Fluticasone propionate | yes | 896 | 36.4 | 83.8 | 14.3 | 96 |
| | no | 6,442 | 36.4 | 83.8 | 14.3 | 96 |
| Fluticasone | yes | 2,154 | 38 | 85.2 | 12.7 | 96.6 |
| | no | 5,184 | 38 | 85.2 | 12.7 | 96.6 |
| *C Streptococcus pneumoniae type X | yes | 510 | 41.9 | 82.4 | 15.7 | 96.9 |
| | no | 6,828 | 41.9 | 82.4 | 15.7 | 96.9 |
| Methylprednisolone acetate | yes | 399 | 34.3 | 86.4 | 11.8 | 96.3 |
| | no | 6,939 | 34.3 | 86.4 | 11.8 | 96.3 |
| Duloxetine | yes | 292 | 25.6 | 90.9 | 8.4 | 98.2 |
| | no | 7,046 | 25.6 | 90.9 | 8.4 | 98.2 |
| Tamsulosin | yes | 216 | 91.5 | 91.8 | 7.6 | 96.6 |
| | no | 7,122 | 91.5 | 91.8 | 7.6 | 96.6 |

| | | | | | | |
|---|---|---|---|---|---|---|
| Naproxen | yes | 982 | 33 | 83.5 | 14.5 | 96 |
| | no | 6,356 | 33 | 83.5 | 14.5 | 96 |
| Folic acid | yes | 417 | 24.1 | 82.8 | 15.2 | 96.8 |
| | no | 6,921 | 24.1 | 82.8 | 15.2 | 96.8 |
| Lutein | yes | 147 | 39.2 | 95.7 | 2.9 | 97.8 |
| | no | 7,191 | 39.2 | 95.7 | 2.9 | 97.8 |
| Fish oils | yes | 437 | 40.9 | 90.1 | 7.4 | 98.1 |
| | no | 6,901 | 40.9 | 90.1 | 7.4 | 98.1 |
| Diclofenac | yes | 605 | 35.9 | 81.1 | 16.7 | 96.3 |
| | no | 6,733 | 35.9 | 81.1 | 16.7 | 96.3 |
| Escitalopram | yes | 499 | 28.9 | 91.1 | 7.9 | 98.5 |
| | no | 6,839 | 28.9 | 91.1 | 7.9 | 98.5 |
| Vitamin B12 | yes | 668 | 28.8 | 85.4 | 12.1 | 97 |
| | no | 6,670 | 28.8 | 85.4 | 12.1 | 97 |
| Codeine | yes | 342 | 25.2 | 90.3 | 9.4 | 98.4 |
| | no | 6,996 | 25.2 | 90.3 | 9.4 | 98.4 |
| Bismuth subsalicylate | yes | 171 | 44.2 | 83.7 | 14.5 | 97.1 |
| | no | 7,167 | 44.2 | 83.7 | 14.5 | 97.1 |
| Amoxicillin | yes | 1,709 | 35.2 | 85.7 | 12 | 96.3 |
| | no | 5,629 | 35.2 | 85.7 | 12 | 96.3 |
| Hydrochlorothiazide | yes | 707 | 41.5 | 79.9 | 18.2 | 98.3 |
| | no | 6,631 | 41.5 | 79.9 | 18.2 | 98.3 |
| Ropivacaine | yes | 219 | 28.1 | 86.3 | 13.3 | 96.8 |
| | no | 7,119 | 28.1 | 86.3 | 13.3 | 96.8 |
| Eicosapentaenoate | yes | 223 | 39.4 | 87.4 | 10 | 97.3 |
| | no | 7,115 | 39.4 | 87.4 | 10 | 97.3 |
| Cefdinir | yes | 298 | 32.1 | 86.2 | 10.2 | 97.9 |
| | no | 7,040 | 32.1 | 86.2 | 10.2 | 97.9 |
| Benzocaine | yes | 413 | 30.2 | 87.9 | 10.3 | 94.4 |
| | no | 6,925 | 30.2 | 87.9 | 10.3 | 94.4 |
| Ketorolac | yes | 551 | 31.3 | 81.3 | 17 | 95.9 |
| | no | 6,787 | 31.3 | 81.3 | 17 | 95.9 |
| Benzonatate | yes | 635 | 32.6 | 85.5 | 12 | 96.1 |
| | no | 6,703 | 32.6 | 85.5 | 12 | 96.1 |
| Cyclobenzaprine | yes | 499 | 27.3 | 82.5 | 16.3 | 96.4 |
| | no | 6,839 | 27.3 | 82.5 | 16.3 | 96.4 |
| Trazodone | yes | 339 | 30.7 | 87.6 | 10.6 | 98.5 |
| | no | 6,999 | 30.7 | 87.6 | 10.6 | 98.5 |
| Zolpidem | yes | 240 | 33.9 | 94.8 | 4.3 | 98.7 |

| | | | | | | |
|---|---|---|---|---|---|---|
| | no | 7,098 | 33.9 | 94.8 | 4.3 | 98.7 |
| Testosterone | yes | 284 | 60.3 | 87.9 | 10.6 | 97.1 |
| | no | 7,054 | 60.3 | 87.9 | 10.6 | 97.1 |
| Simvastatin | yes | 203 | 52.1 | 91.2 | 7.3 | 97.7 |
| | no | 7,135 | 52.1 | 91.2 | 7.3 | 97.7 |
| Cephalexin | yes | 416 | 33.2 | 88.3 | 10.5 | 97.8 |
| | no | 6,922 | 33.2 | 88.3 | 10.5 | 97.8 |
| Varicella zoster virus glycoprotein E | yes | 222 | 39.6 | 84.7 | 11.5 | 98.6 |
| | no | 7,116 | 39.6 | 84.7 | 11.5 | 98.6 |
| Doxycycline | yes | 693 | 36.4 | 87.9 | 10.7 | 97.6 |
| | no | 6,645 | 36.4 | 87.9 | 10.7 | 97.6 |
| Diclofenac sodium | yes | 341 | 30.6 | 82.1 | 15.6 | 97.3 |
| | no | 6,997 | 30.6 | 82.1 | 15.6 | 97.3 |
| Butalbital | yes | 347 | 28.2 | 81.2 | 15.7 | 97.1 |
| | no | 6,991 | 28.2 | 81.2 | 15.7 | 97.1 |
| Propofol | yes | 602 | 42.5 | 84.7 | 12.8 | 96.8 |
| | no | 6,736 | 42.5 | 84.7 | 12.8 | 96.8 |
| Cetirizine | yes | 1,388 | 37.5 | 83.6 | 13.6 | 96 |
| | no | 5,950 | 37.5 | 83.6 | 13.6 | 96 |
| Oxymetazoline | yes | 1,143 | 39.6 | 85.9 | 11.5 | 96.3 |
| | no | 6,195 | 39.6 | 85.9 | 11.5 | 96.3 |
| Diethylstilbestrol | yes | 172 | 43.4 | 91.6 | 6.7 | 97 |
| | no | 7,166 | 43.4 | 91.6 | 6.7 | 97 |

Other abbreviations and acronyms. *A Streptococcus pneumoniae serotype X: Streptococcus pneumoniae serotype (1, 19A, 3, 5, 6A, 7F) capsular antigen diphtheria CRM197 protein conjugate vaccine; *B Streptococcus pneumoniae serotype X: Streptococcus pneumoniae serotype (14, 18C, 19F, 23F, 4, 6B, 9V) capsular antigen diphtheria CRM197 protein conjugate vaccine; *C Streptococcus pneumoniae type X: Streptococcus pneumoniae type (1, 10A, 11A, 12F, 14, 15B, 17F, 18C, 19A, 19F, 2, 20, 22F, 23F, 3, 33F, 4, 5, 6B, 7F, 8, 9N, 9V) capsular polysaccharide antigen.

**eTable 5** Association between drug exposure and on ventilator (cumulative severity).

**TCe/u**, total count of drug exposed/unexposed patients; **SRe/u**, severity rate of drug exposed/unexposed patients computed after overlap weighting with propensity score; **OR**, odds ratio; **CI**, confidence interval. The association results are arranged in the ascending order of OR values. The statistically significant results at the 0.05 level are emphasized in bold.

| Drug name | Drug class | TCe | TCu | SRe | SRu | OR | 95% CI |
|---|---|---|---|---|---|---|---|
| Clavulanate | antibiotic | 867 | 6,495 | 0.8 | 2 | 0.4 | (0.15 - 1.07) |
| ***A Streptococcus pneumoniae serotype X** | **vaccine** | **390** | **6,972** | **2.6** | **4.7** | **0.41** | **(0.17 - 0.99)** |
| Sildenafil | PDE5 inhibitor | 180 | 7,182 | 2.6 | 6.9 | 0.42 | (0.12 - 1.41) |
| Flaxseed extract | supplement | 188 | 7,174 | 3.8 | 9.2 | 0.44 | (0.16 - 1.24) |
| Ascorbic acid | supplement | 566 | 6,796 | 1.4 | 2.9 | 0.46 | (0.18 - 1.17) |
| Loratadine | antihistamine | 756 | 6,606 | 0.9 | 1.9 | 0.47 | (0.17 - 1.29) |
| ***B Streptococcus pneumoniae serotype X** | **vaccine** | **617** | **6,745** | **2.5** | **4.1** | **0.47** | **(0.22 - 0.99)** |
| Diphtheria toxoid vaccine, inactivated | vaccine | 1,171 | 6,191 | 1.1 | 2 | 0.51 | (0.23 - 1.09) |
| Tetanus toxoid vaccine, inactivated | vaccine | 1,174 | 6,188 | 1.1 | 2 | 0.51 | (0.23 - 1.10) |
| Acellular pertussis vaccine, inactivated | vaccine | 1,124 | 6,238 | 1.2 | 2 | 0.53 | (0.24 - 1.15) |
| Methylprednisolone acetate | corticosteroid | 399 | 6,963 | 1.2 | 2.8 | 0.54 | (0.17 - 1.76) |
| Omega-3 fatty acids | supplement | 436 | 6,926 | 3.3 | 5.1 | 0.59 | (0.27 - 1.27) |
| Meloxicam | NSAID | 591 | 6,771 | 1.5 | 3.2 | 0.61 | (0.25 - 1.50) |
| Pseudoephedrine | decongestant | 952 | 6,410 | 0.6 | 1.5 | 0.61 | (0.21 - 1.79) |
| Fluticasone | corticosteroid | 2,161 | 5,201 | 1.3 | 2.1 | 0.64 | (0.35 - 1.16) |
| Bismuth subsalicylate | antidiarrheal | 171 | 7,191 | 2.8 | 3.6 | 0.65 | (0.17 - 2.48) |
| Naproxen | NSAID | 984 | 6,378 | 1.1 | 2 | 0.66 | (0.28 - 1.54) |
| Ibuprofen | NSAID | 3,518 | 3,844 | 1.1 | 1.7 | 0.66 | (0.37 - 1.20) |
| Cefdinir | antibiotic | 298 | 7,064 | 1.7 | 1.4 | 0.68 | (0.15 - 3.20) |
| *C Streptococcus pneumoniae type X | vaccine | 515 | 6,847 | 3.5 | 5 | 0.71 | (0.35 - 1.45) |
| Fluticasone propionate | corticosteroid | 900 | 6,462 | 1.4 | 2 | 0.72 | (0.32 - 1.62) |
| Vitamin B12 | supplement | 670 | 6,692 | 2.4 | 2.9 | 0.73 | (0.34 - 1.60) |
| Varicella zoster virus glycoprotein E | vaccine | 222 | 7,140 | 2.9 | 6.1 | 0.75 | (0.24 - 2.37) |
| Benzonatate | antitussive | 636 | 6,726 | 2 | 2.8 | 0.77 | (0.34 - 1.75) |
| Tamsulosin | alpha 1 blocker | 219 | 7,143 | 5.5 | 6.9 | 0.79 | (0.32 - 1.94) |
| Escitalopram | antidepressant | 500 | 6,862 | 1.4 | 1.4 | 0.8 | (0.24 - 2.67) |
| Azelastine | antihistamine | 482 | 6,880 | 2 | 2.8 | 0.81 | (0.32 - 2.04) |
| Amoxicillin | antibiotic | 1,713 | 5,649 | 1.3 | 1.4 | 0.81 | (0.41 - 1.62) |
| Duloxetine | antidepressant | 294 | 7,068 | 2.3 | 3 | 0.81 | (0.27 - 2.42) |
| Ranitidine | antacid | 252 | 7,110 | 1.9 | 2.4 | 0.81 | (0.22 - 3.00) |
| Tetracaine | anesthetic | 200 | 7,162 | 2.5 | 2.6 | 0.82 | (0.21 - 3.15) |
| Cetirizine | antihistamine | 1,390 | 5,972 | 1.3 | 1.6 | 0.84 | (0.40 - 1.74) |
| Ropivacaine | anesthetic | 220 | 7,142 | 2.7 | 3.6 | 0.84 | (0.26 - 2.75) |
| Valacyclovir | antiviral | 453 | 6,909 | 1.3 | 2 | 0.85 | (0.26 - 2.70) |

| | | | | | | | |
|---|---|---|---|---|---|---|---|
| Lutein | supplement | 149 | 7,213 | 5.6 | 7.3 | 0.86 | (0.30 - 2.44) |
| Ketorolac | NSAID | 553 | 6,809 | 1.4 | 2 | 0.86 | (0.31 - 2.38) |
| Zolpidem | hypnotic | 241 | 7,121 | 2.4 | 3.8 | 0.88 | (0.27 - 2.88) |
| Diclofenac sodium | NSAID | 342 | 7,020 | 3.3 | 4.2 | 0.89 | (0.37 - 2.12) |
| Diclofenac | NSAID | 610 | 6,752 | 2.6 | 3.4 | 0.9 | (0.42 - 1.90) |
| Hydrochlorothiazide | antihypertensive | 714 | 6,648 | 3.5 | 4.8 | 0.91 | (0.48 - 1.70) |
| Ketoconazole | antifungal | 230 | 7,132 | 2.1 | 2.1 | 0.94 | (0.24 - 3.79) |
| Butalbital | barbituate | 348 | 7,014 | 2 | 2.3 | 0.95 | (0.32 - 2.84) |
| Losartan | antihypertensive | 464 | 6,898 | 3.8 | 5 | 0.95 | (0.46 - 1.98) |
| Fish oils | supplement | 442 | 6,920 | 3.5 | 3.8 | 0.97 | (0.45 - 2.11) |
| Tropicamide | opthalmic agent | 352 | 7,010 | 3.2 | 3.2 | 0.97 | (0.38 - 2.47) |
| Eicosapentaenoate | supplement | 225 | 7,137 | 3.5 | 4 | 0.98 | (0.34 - 2.83) |
| Trazodone | antidepressant | 341 | 7,021 | 2.8 | 2.8 | 0.99 | (0.36 - 2.71) |
| Methylprednisolone | corticosteroid | 876 | 6,486 | 2.1 | 2.5 | 1 | (0.48 - 2.04) |

Other abbreviations and acronyms. *A Streptococcus pneumoniae serotype X: Streptococcus pneumoniae serotype (1, 19A, 3, 5, 6A, 7F) capsular antigen diphtheria CRM197 protein conjugate vaccine; *B Streptococcus pneumoniae serotype X: Streptococcus pneumoniae serotype (14, 18C, 19F, 23F, 4, 6B, 9V) capsular antigen diphtheria CRM197 protein conjugate vaccine; *C Streptococcus pneumoniae type X: Streptococcus pneumoniae type (1, 10A, 11A, 12F, 14, 15B, 17F, 18C, 19A, 19F, 2, 20, 22F, 23F, 3, 33F, 4, 5, 6B, 7F, 8, 9N, 9V) capsular polysaccharide antigen; NSAID: nonsteroidal anti-inflammatory drug; PDE5 inhibitor: phosphodiesterase type 5 inhibitor.

**eTable 6** Association between drug exposure and in ICU (cumulative severity).

**TCe/u**, total count of drug exposed/unexposed patients; **SRe/u**, severity rate of drug exposed/unexposed patients computed after overlap weighting with propensity score; **OR**, odds ratio; **CI**, confidence interval. The association results are arranged in the ascending order of OR values. The statistically significant results at the 0.05 level are emphasized in bold.

| Drug name | Drug class | TCe | TCu | SRe | SRu | OR | 95% CI |
|---|---|---|---|---|---|---|---|
| ***A Streptococcus pneumoniae serotype X** | **vaccine** | **391** | **7,001** | **2.8** | **5.4** | **0.39** | **(0.17 - 0.89)** |
| ***B Streptococcus pneumoniae serotype X** | **vaccine** | **618** | **6,774** | **2.7** | **4.9** | **0.41** | **(0.20 - 0.83)** |
| **Diphtheria toxoid vaccine, inactivated** | **vaccine** | **1,172** | **6,220** | **1.2** | **2.6** | **0.42** | **(0.21 - 0.87)** |
| **Tetanus toxoid vaccine, inactivated** | **vaccine** | **1,175** | **6,217** | **1.2** | **2.6** | **0.42** | **(0.21 - 0.87)** |
| **Acellular pertussis vaccine, inactivated** | **vaccine** | **1,125** | **6,267** | **1.3** | **2.5** | **0.44** | **(0.21 - 0.91)** |
| Flaxseed extract | supplement | 189 | 7,203 | 4.3 | 9.9 | 0.44 | (0.17 - 1.18) |
| Dextromethorphan | antitussive | 900 | 6,492 | 0.8 | 2 | 0.45 | (0.17 - 1.17) |
| Sildenafil | PDE5 inhibitor | 182 | 7,210 | 3.7 | 7.6 | 0.51 | (0.18 - 1.45) |
| Fexofenadine | antihistamine | 560 | 6,832 | 1.2 | 2.6 | 0.52 | (0.19 - 1.38) |
| Methylprednisolone acetate | corticosteroid | 400 | 6,992 | 1.5 | 3.4 | 0.52 | (0.18 - 1.51) |
| Naproxen | NSAID | 985 | 6,407 | 1.2 | 2.5 | 0.55 | (0.25 - 1.20) |
| Clavulanate | antibiotic | 872 | 6,520 | 1.4 | 2.4 | 0.57 | (0.26 - 1.25) |
| Pseudoephedrine | decongestant | 954 | 6,438 | 0.8 | 1.9 | 0.6 | (0.24 - 1.54) |
| *C Streptococcus pneumoniae type X | vaccine | 516 | 6,876 | 3.7 | 5.9 | 0.61 | (0.31 - 1.20) |
| Omega-3 fatty acids | supplement | 439 | 6,953 | 4 | 5.7 | 0.64 | (0.32 - 1.30) |
| Ascorbic acid | supplement | 571 | 6,821 | 2.3 | 3.4 | 0.65 | (0.30 - 1.41) |
| Bismuth subsalicylate | antidiarrheal | 172 | 7,220 | 3.4 | 4.2 | 0.65 | (0.19 - 2.18) |
| Fluticasone | corticosteroid | 2,169 | 5,223 | 1.6 | 2.6 | 0.65 | (0.39 - 1.11) |
| Varicella zoster virus glycoprotein E | vaccine | 223 | 7,169 | 3.3 | 6.8 | 0.66 | (0.23 - 1.89) |
| Fluticasone propionate | corticosteroid | 902 | 6,490 | 1.6 | 2.5 | 0.66 | (0.32 - 1.39) |
| Loratadine | antihistamine | 761 | 6,631 | 1.6 | 2.3 | 0.68 | (0.30 - 1.54) |
| Meloxicam | NSAID | 595 | 6,797 | 2.1 | 3.7 | 0.69 | (0.32 - 1.49) |
| Betamethasone | corticosteroid | 238 | 7,154 | 2.5 | 3.6 | 0.72 | (0.23 - 2.24) |
| Escitalopram | antidepressant | 501 | 6,891 | 1.6 | 1.8 | 0.72 | (0.24 - 2.13) |
| Ibuprofen | NSAID | 3,531 | 3,861 | 1.5 | 2.1 | 0.73 | (0.44 - 1.22) |
| Ketoconazole | antifungal | 230 | 7,162 | 2.1 | 2.5 | 0.76 | (0.20 - 2.82) |
| Diclofenac sodium | NSAID | 343 | 7,049 | 3.6 | 4.9 | 0.8 | (0.35 - 1.80) |
| Azelastine | antihistamine | 484 | 6,908 | 2.4 | 3.3 | 0.8 | (0.34 - 1.85) |
| Ropivacaine | anesthetic | 221 | 7,171 | 3.2 | 4.2 | 0.81 | (0.28 - 2.39) |
| Codeine | opioid | 345 | 7,047 | 2.7 | 4 | 0.81 | (0.32 - 2.08) |
| Eicosapentaenoate | supplement | 225 | 7,167 | 3.5 | 4.6 | 0.82 | (0.30 - 2.27) |
| Tetracaine | anesthetic | 201 | 7,191 | 2.9 | 3.2 | 0.83 | (0.24 - 2.85) |
| Zolpidem | hypnotic | 243 | 7,149 | 3.2 | 4.4 | 0.84 | (0.29 - 2.40) |
| Lutein | supplement | 150 | 7,242 | 6.3 | 7.9 | 0.85 | (0.32 - 2.25) |

| | | | | | | | |
|---|---|---|---|---|---|---|---|
| Duloxetine | antidepressant | 296 | 7,096 | 2.9 | 3.5 | 0.85 | (0.32 - 2.28) |
| Butalbital | barbituate | 349 | 7,043 | 2.2 | 2.8 | 0.86 | (0.31 - 2.37) |
| Vitamin B12 | supplement | 676 | 6,716 | 3.3 | 3.5 | 0.88 | (0.45 - 1.72) |
| Tamsulosin | alpha 1 blocker | 222 | 7,170 | 6.7 | 7.5 | 0.88 | (0.38 - 2.03) |
| Valacyclovir | antiviral | 455 | 6,937 | 1.7 | 2.4 | 0.88 | (0.32 - 2.43) |
| Amoxicillin | antibiotic | 1,721 | 5,671 | 1.7 | 1.8 | 0.89 | (0.48 - 1.62) |
| Folic acid | supplement | 422 | 6,970 | 3 | 2.8 | 0.89 | (0.37 - 2.15) |
| Diclofenac | NSAID | 614 | 6,778 | 3.2 | 4.1 | 0.9 | (0.46 - 1.75) |
| Benzonatate | antitussive | 641 | 6,751 | 2.7 | 3.3 | 0.9 | (0.44 - 1.85) |
| Cetirizine | antihistamine | 1,396 | 5,996 | 1.7 | 2 | 0.9 | (0.47 - 1.71) |
| Testosterone | androgen | 287 | 7,105 | 3.1 | 3.4 | 0.91 | (0.33 - 2.48) |
| Methylprednisolone | corticosteroid | 879 | 6,513 | 2.4 | 3 | 0.91 | (0.47 - 1.76) |
| Phenol | anesthetic | 339 | 7,053 | 1.5 | 1.6 | 0.91 | (0.25 - 3.29) |
| Losartan | antihypertensive | 467 | 6,925 | 4.4 | 5.8 | 0.91 | (0.47 - 1.80) |
| Ketorolac | NSAID | 556 | 6,836 | 1.9 | 2.5 | 0.92 | (0.38 - 2.21) |
| Alprazolam | benzodiazepine | 334 | 7,058 | 2.4 | 2.8 | 0.93 | (0.31 - 2.74) |
| Cefdinir | antibiotic | 300 | 7,092 | 2.3 | 1.7 | 0.93 | (0.26 - 3.35) |
| Tropicamide | opthalmic agent | 354 | 7,038 | 3.8 | 3.7 | 0.95 | (0.41 - 2.23) |
| Adenosine monophosphate | antiarrhythmic agent | 178 | 7,214 | 4.2 | 4.5 | 0.97 | (0.30 - 3.11) |
| Ranitidine | antacid | 254 | 7,138 | 2.7 | 3 | 0.97 | (0.32 - 3.00) |
| Menthol | topical skin product | 366 | 7,026 | 3.2 | 2.8 | 0.98 | (0.39 - 2.45) |
| Docosahexaenoate | supplement | 171 | 7,221 | 2.9 | 2.9 | 0.99 | (0.26 - 3.74) |
| Diethylstilbestrol | estrogen | 175 | 7,217 | 6.1 | 7.1 | 0.99 | (0.38 - 2.58) |
| Fish oils | supplement | 445 | 6,947 | 4.1 | 4.3 | 0.99 | (0.49 - 2.03) |
| Pamidronate | bisphosphonate derivative | 244 | 7,148 | 4.7 | 4.1 | 1 | (0.39 - 2.56) |
| Trazodone | antidepressant | 343 | 7,049 | 3.4 | 3.3 | 1 | (0.40 - 2.47) |
| Esomeprazole | antacid | 172 | 7,220 | 3.4 | 4.2 | 1 | (0.30 - 3.37) |

Other abbreviations and acronyms. *A Streptococcus pneumoniae serotype X: Streptococcus pneumoniae serotype (1, 19A, 3, 5, 6A, 7F) capsular antigen diphtheria CRM197 protein conjugate vaccine; *B Streptococcus pneumoniae serotype X: Streptococcus pneumoniae serotype (14, 18C, 19F, 23F, 4, 6B, 9V) capsular antigen diphtheria CRM197 protein conjugate vaccine; *C Streptococcus pneumoniae type X: Streptococcus pneumoniae type (1, 10A, 11A, 12F, 14, 15B, 17F, 18C, 19A, 19F, 2, 20, 22F, 23F, 3, 33F, 4, 5, 6B, 7F, 8, 9N, 9V) capsular polysaccharide antigen; NSAID: nonsteroidal anti-inflammatory drug; PDE5 inhibitor: phosphodiesterase type 5 inhibitor.

**eTable 7** Association between drug exposure and hospitalized-mild (cumulative severity).

**TCe/u**, total count of drug exposed/unexposed patients; **SRe/u**, severity rate of drug exposed/unexposed patients computed after overlap weighting with propensity score; **OR**, odds ratio; **CI**, confidence interval. The association results are arranged in the ascending order of OR values. The statistically significant results at the 0.05 level are emphasized in bold.

| Drug name | Drug class | TCe | TCu | SRe | SRu | OR | 95% CI |
|---|---|---|---|---|---|---|---|
| **Ethinyl estradiol** | **estrogen** | **749** | **7,019** | **1** | **2.8** | **0.37** | **(0.14 - 0.99)** |
| Clotrimazole | antifungal | 160 | 7,608 | 4.3 | 9 | 0.4 | (0.14 - 1.15) |
| **Turmeric extract** | **supplement** | **154** | **7,614** | **5.8** | **14.2** | **0.4** | **(0.17 - 0.98)** |
| Triamcinolone acetonide | corticosteroid | 154 | 7,614 | 5.1 | 11.2 | 0.41 | (0.15 - 1.07) |
| **Flaxseed extract** | **supplement** | **206** | **7,562** | **11.6** | **23.6** | **0.41** | **(0.22 - 0.77)** |
| Levonorgestrel | estrogen | 507 | 7,261 | 1.2 | 3.4 | 0.42 | (0.15 - 1.18) |
| Ubidecarenone | supplement | 131 | 7,637 | 7.5 | 16.3 | 0.44 | (0.19 - 1.02) |
| ***C Streptococcus pneumoniae type X** | **vaccine** | **552** | **7,216** | **9.4** | **16.8** | **0.47** | **(0.31 - 0.72)** |
| **Methylprednisolone acetate** | **corticosteroid** | **413** | **7,355** | **4.5** | **10** | **0.48** | **(0.26 - 0.89)** |
| ***A Streptococcus pneumoniae serotype X** | **vaccine** | **419** | **7,349** | **9.1** | **14.9** | **0.49** | **(0.30 - 0.80)** |
| **Estradiol** | **estrogen derivative** | **576** | **7,192** | **3.1** | **6.2** | **0.51** | **(0.26 - 0.97)** |
| Biotin | supplement | 209 | 7,559 | 7.2 | 12.3 | 0.51 | (0.24 - 1.09) |
| Estrogens | estrogen | 316 | 7,452 | 4.4 | 8.5 | 0.53 | (0.26 - 1.09) |
| ***B Streptococcus pneumoniae serotype X** | **vaccine** | **667** | **7,101** | **9.4** | **14.1** | **0.55** | **(0.37 - 0.82)** |
| Oseltamivir | antiviral | 557 | 7,211 | 3.4 | 6 | 0.55 | (0.29 - 1.04) |
| **Pseudoephedrine** | **decongestant** | **978** | **6,790** | **3.3** | **6.3** | **0.58** | **(0.35 - 0.94)** |
| **Omega-3 fatty acids** | **supplement** | **475** | **7,293** | **10.7** | **15.7** | **0.6** | **(0.39 - 0.94)** |
| Psyllium | antidiarrheal | 116 | 7,652 | 6.8 | 11.4 | 0.61 | (0.22 - 1.67) |
| Eicosapentaenoate | supplement | 237 | 7,531 | 8.2 | 13 | 0.61 | (0.32 - 1.18) |
| Glucosamine | supplement | 113 | 7,655 | 8.7 | 15.2 | 0.62 | (0.25 - 1.55) |
| Haemophilus influenzae type b | vaccine | 373 | 7,395 | 4.5 | 6.7 | 0.64 | (0.32 - 1.27) |
| Varicella zoster virus glycoprotein E | vaccine | 241 | 7,527 | 10.3 | 17.6 | 0.64 | (0.35 - 1.19) |
| Betamethasone | corticosteroid | 250 | 7,518 | 7.1 | 10.6 | 0.65 | (0.33 - 1.29) |
| Azelastine | antihistamine | 507 | 7,261 | 6.7 | 10.1 | 0.66 | (0.40 - 1.10) |
| Dextromethorphan | antitussive | 933 | 6,835 | 4.3 | 6.6 | 0.66 | (0.42 - 1.06) |
| **Tetanus toxoid vaccine, inactivated** | **vaccine** | **1,233** | **6,535** | **5.8** | **8** | **0.67** | **(0.47 - 0.97)** |
| Estrogens, conjugated (USP) | estrogens | 121 | 7,647 | 8.3 | 12.7 | 0.67 | (0.26 - 1.72) |
| Fexofenadine | antihistamine | 586 | 7,182 | 5.5 | 8.3 | 0.67 | (0.40 - 1.12) |
| **Ibuprofen** | **NSAID** | **3,655** | **4,113** | **5.2** | **7.4** | **0.68** | **(0.51 - 0.90)** |
| **Fluticasone** | **corticosteroid** | **2,271** | **5,497** | **5.7** | **8.3** | **0.68** | **(0.50 - 0.91)** |
| **Diphtheria toxoid vaccine, inactivated** | **vaccine** | **1,231** | **6,537** | **5.9** | **8** | **0.68** | **(0.48 - 0.98)** |
| Calcium citrate | calcium salt | 116 | 7,652 | 7.8 | 12 | 0.68 | (0.27 - 1.75) |
| **Acellular pertussis vaccine, inactivated** | **vaccine** | **1,181** | **6,587** | **5.9** | **8** | **0.69** | **(0.47 - 0.99)** |
| Naproxen | NSAID | 1,029 | 6,739 | 5.3 | 8 | 0.69 | (0.46 - 1.03) |

| | | | | | | | |
|---|---|---|---|---|---|---|---|
| Tadalafil | PDE5 inhibitor | 131 | 7,637 | 12.3 | 18.1 | 0.71 | (0.34 - 1.50) |
| Clobetasol | corticosteroid | 156 | 7,612 | 8.2 | 10.5 | 0.71 | (0.31 - 1.66) |
| Follicle stimulating hormone | gonadotropin | 134 | 7,634 | 4.5 | 6.6 | 0.73 | (0.23 - 2.30) |
| Nitrofurantoin, monohydrate | antibiotic | 210 | 7,558 | 4.8 | 6.6 | 0.73 | (0.28 - 1.87) |
| Cetirizine | antihistamine | 1,447 | 6,321 | 5.1 | 7 | 0.73 | (0.51 - 1.05) |
| Fluticasone propionate | corticosteroid | 947 | 6,821 | 6.2 | 8 | 0.73 | (0.49 - 1.10) |
| Phentermine | anorexiant | 123 | 7,645 | 5 | 6.5 | 0.74 | (0.22 - 2.47) |
| Sildenafil | PDE5 inhibitor | 203 | 7,565 | 13.8 | 18.3 | 0.76 | (0.41 - 1.39) |
| Loratadine | antihistamine | 797 | 6,971 | 5.9 | 7.7 | 0.76 | (0.48 - 1.18) |
| Lutein | supplement | 167 | 7,601 | 15.9 | 20 | 0.78 | (0.41 - 1.46) |
| Adalimumab | DMARD | 132 | 7,636 | 6 | 8.4 | 0.78 | (0.28 - 2.15) |
| Lamotrigine | anticonvulsant | 160 | 7,608 | 5 | 6.1 | 0.78 | (0.27 - 2.24) |
| Vitamin B12 | supplement | 728 | 7,040 | 9.6 | 11.3 | 0.8 | (0.54 - 1.19) |
| Naproxen sodium | NSAID | 137 | 7,631 | 7.9 | 10 | 0.8 | (0.33 - 1.97) |
| Testosterone | androgen | 302 | 7,466 | 7.8 | 9.8 | 0.81 | (0.43 - 1.51) |
| Benzonatate | antitussive | 680 | 7,088 | 8.1 | 10.1 | 0.82 | (0.53 - 1.24) |
| Cortisone | corticosteroid | 179 | 7,589 | 9.3 | 11.9 | 0.82 | (0.39 - 1.72) |
| Diethylstilbestrol | estrogen | 193 | 7,575 | 14.7 | 18.1 | 0.82 | (0.45 - 1.50) |
| Diclofenac sodium | NSAID | 375 | 7,393 | 11.4 | 13.9 | 0.85 | (0.53 - 1.39) |
| Lufenuron | antiparasitic/antifungal | 217 | 7,551 | 9.5 | 10.3 | 0.87 | (0.44 - 1.71) |
| Oxymetazoline | adrenergic agonist agent | 1,206 | 6,562 | 6.6 | 7.1 | 0.87 | (0.61 - 1.26) |
| Fish oils | supplement | 479 | 7,289 | 10.7 | 12.4 | 0.88 | (0.56 - 1.38) |
| Meloxicam | NSAID | 640 | 7,128 | 8.7 | 10.7 | 0.9 | (0.59 - 1.37) |
| Finasteride | 5-alpha reductase inhibitor | 106 | 7,662 | 13.8 | 15 | 0.91 | (0.39 - 2.09) |
| Ascorbic acid | supplement | 618 | 7,150 | 9.5 | 10.3 | 0.91 | (0.59 - 1.38) |
| Desoxycorticosterone | corticosteroid | 159 | 7,609 | 11.2 | 12 | 0.92 | (0.43 - 1.96) |
| Tretinoin | antineoplastic agent | 230 | 7,538 | 3.5 | 4.4 | 0.92 | (0.33 - 2.58) |
| Ketoconazole | antifungal | 244 | 7,524 | 7.7 | 8 | 0.93 | (0.45 - 1.89) |
| Norethindrone | contraceptive | 441 | 7,327 | 2.6 | 3.1 | 0.93 | (0.38 - 2.28) |
| Methylprednisolone | corticosteroid | 933 | 6,835 | 7.9 | 9.3 | 0.93 | (0.64 - 1.35) |
| Varicella-zoster virus vaccine live (Oka-Merck) strain | vaccine | 165 | 7,603 | 9.7 | 8.4 | 0.94 | (0.41 - 2.14) |
| Fiber | supplement | 699 | 7,069 | 7.4 | 8.3 | 0.94 | (0.61 - 1.45) |
| Docosahexaenoate | supplement | 181 | 7,587 | 8.3 | 8.9 | 0.94 | (0.43 - 2.05) |
| Thioguanine | antineoplastic agent | 252 | 7,516 | 12.4 | 13.6 | 0.94 | (0.53 - 1.68) |
| Celecoxib | NSAID | 182 | 7,586 | 11.4 | 12.9 | 0.94 | (0.47 - 1.89) |
| Mometasone | corticosteroid | 222 | 7,546 | 8.5 | 9.6 | 0.95 | (0.46 - 1.95) |
| Diclofenac | NSAID | 667 | 7,101 | 10.5 | 11.8 | 0.95 | (0.65 - 1.40) |
| Phenol | anesthetic | 354 | 7,414 | 5.7 | 5.8 | 0.97 | (0.49 - 1.92) |
| Metoprolol succinate | antihypertensive | 195 | 7,573 | 16.3 | 17.3 | 0.98 | (0.54 - 1.77) |

| | | | | | | | |
|---|---|---|---|---|---|---|---|
| Hepatitis A vaccine (inactivated) strain HM175 | vaccine | 189 | 7,579 | 7.4 | 6.8 | 0.99 | (0.41 - 2.35) |
| Hepatitis A Vaccine, Inactivated | vaccine | 166 | 7,602 | 6 | 5.8 | 0.99 | (0.36 - 2.73) |
| Valacyclovir | antiviral | 482 | 7,286 | 7.2 | 8 | 1 | (0.58 - 1.71) |
| Guaifenesin | expectorant | 1,293 | 6,475 | 8.3 | 8.1 | 1 | (0.71 - 1.40) |

Other abbreviations and acronyms. *A Streptococcus pneumoniae serotype X: Streptococcus pneumoniae serotype (1, 19A, 3, 5, 6A, 7F) capsular antigen diphtheria CRM197 protein conjugate vaccine; *B Streptococcus pneumoniae serotype X: Streptococcus pneumoniae serotype (14, 18C, 19F, 23F, 4, 6B, 9V) capsular antigen diphtheria CRM197 protein conjugate vaccine; *C Streptococcus pneumoniae type X: Streptococcus pneumoniae type (1, 10A, 11A, 12F, 14, 15B, 17F, 18C, 19A, 19F, 2, 20, 22F, 23F, 3, 33F, 4, 5, 6B, 7F, 8, 9N, 9V) capsular polysaccharide antigen; NSAID: nonsteroidal anti-inflammatory drug; PDE5 inhibitor: phosphodiesterase type 5 inhibitor; DMARD: disease-modifying antirheumatic drugs.

**eTable 8** Association between drug exposure and on ventilator (exclusive severity).

**TCe/u**, total count of drug exposed/unexposed patients; **SRe/u**, severity rate of drug exposed/unexposed patients computed after overlap weighting with propensity score; **OR**, odds ratio; **CI**, confidence interval. The association results are arranged in the ascending order of OR values.

| Drug name | Drug class | TCe | TCu | SRe | SRu | OR | 95% CI |
|---|---|---|---|---|---|---|---|
| *A Streptococcus pneumoniae serotype X | vaccine | 384 | 6,928 | 1.3 | 2.1 | 0.52 | (0.15 - 1.81) |
| Tetanus toxoid vaccine, inactivated | vaccine | 1,168 | 6,144 | 0.7 | 1.1 | 0.53 | (0.19 - 1.44) |
| Diphtheria toxoid vaccine, inactivated | vaccine | 1,165 | 6,147 | 0.7 | 1.1 | 0.53 | (0.19 - 1.44) |
| Acellular pertussis vaccine, inactivated | vaccine | 1,118 | 6,194 | 0.7 | 1.1 | 0.55 | (0.20 - 1.52) |
| Ascorbic acid | supplement | 562 | 6,750 | 0.8 | 1.4 | 0.59 | (0.17 - 2.03) |
| Vitamin B12 | supplement | 657 | 6,655 | 0.9 | 1.4 | 0.6 | (0.19 - 1.86) |
| *B Streptococcus pneumoniae serotype X | vaccine | 608 | 6,704 | 1.4 | 1.9 | 0.62 | (0.23 - 1.69) |
| Fluticasone | corticosteroid | 2,147 | 5,165 | 0.7 | 1.2 | 0.65 | (0.30 - 1.41) |
| Ibuprofen | NSAID | 3,505 | 3,807 | 0.7 | 1 | 0.69 | (0.32 - 1.46) |
| Cetirizine | antihistamine | 1,381 | 5,931 | 0.6 | 0.9 | 0.7 | (0.27 - 1.85) |
| Amlodipine | antihypertensive | 474 | 6,838 | 1.8 | 2.7 | 0.73 | (0.27 - 1.93) |
| Losartan | antihypertensive | 451 | 6,861 | 1.6 | 2.4 | 0.73 | (0.26 - 2.07) |
| Amoxicillin | antibiotic | 1,703 | 5,609 | 0.7 | 0.9 | 0.74 | (0.30 - 1.84) |
| Fluticasone propionate | corticosteroid | 894 | 6,418 | 0.8 | 1.1 | 0.74 | (0.25 - 2.18) |
| Sildenafil | PDE5 inhibitor | 180 | 7,132 | 2.6 | 3.4 | 0.81 | (0.21 - 3.16) |
| Benzonatate | antitussive | 630 | 6,682 | 1.1 | 1.3 | 0.82 | (0.27 - 2.45) |
| Tamsulosin | alpha 1 blocker | 212 | 7,100 | 3.2 | 3.7 | 0.85 | (0.26 - 2.73) |
| Naproxen | NSAID | 981 | 6,331 | 0.8 | 1 | 0.88 | (0.31 - 2.47) |
| Omega-3 fatty acids | supplement | 429 | 6,883 | 2.1 | 2.2 | 0.89 | (0.32 - 2.47) |
| Ketorolac | NSAID | 550 | 6,762 | 0.9 | 1.1 | 0.89 | (0.25 - 3.22) |
| Tropicamide | opthalmic agent | 345 | 6,967 | 1.4 | 1.5 | 0.9 | (0.24 - 3.43) |
| Meloxicam | NSAID | 589 | 6,723 | 1.1 | 1.4 | 0.94 | (0.31 - 2.86) |
| Dexamethasone | corticosteroid | 880 | 6,432 | 1 | 1.1 | 0.97 | (0.36 - 2.62) |
| *C Streptococcus pneumoniae type X | vaccine | 505 | 6,807 | 2.1 | 2.1 | 0.99 | (0.38 - 2.60) |

Other abbreviations and acronyms. *A Streptococcus pneumoniae serotype X: Streptococcus pneumoniae serotype (1, 19A, 3, 5, 6A, 7F) capsular antigen diphtheria CRM197 protein conjugate vaccine; *B Streptococcus pneumoniae serotype X: Streptococcus pneumoniae serotype (14, 18C, 19F, 23F, 4, 6B, 9V) capsular antigen diphtheria CRM197 protein conjugate vaccine; *C Streptococcus pneumoniae type X: Streptococcus pneumoniae type (1, 10A, 11A, 12F, 14, 15B, 17F, 18C, 19A, 19F, 2, 20, 22F, 23F, 3, 33F, 4, 5, 6B, 7F, 8, 9N, 9V) capsular polysaccharide antigen; NSAID: nonsteroidal anti-inflammatory drug; PDE5 inhibitor: phosphodiesterase type 5 inhibitor.

**eTable 9** Association between drug exposure and in ICU (exclusive severity).

**TCe/u**, total count of drug exposed/unexposed patients; **SRe/u**, severity rate of drug exposed/unexposed patients computed after overlap weighting with propensity score; **OR**, odds ratio; **CI**, confidence interval. The association results are arranged in the ascending order of OR values. The statistically significant results at the 0.05 level are emphasized in bold.

| Drug name | Drug class | TCe | TCu | SRe | SRu | OR | 95% CI |
|---|---|---|---|---|---|---|---|
| **Diphtheria toxoid vaccine, inactivated** | **vaccine** | **1,164** | **6,166** | **0.6** | **1.5** | **0.34** | **(0.12 - 0.93)** |
| **Tetanus toxoid vaccine, inactivated** | **vaccine** | **1,167** | **6,163** | **0.6** | **1.5** | **0.34** | **(0.12 - 0.93)** |
| **Acellular pertussis vaccine, inactivated** | **vaccine** | **1,117** | **6,213** | **0.6** | **1.5** | **0.35** | **(0.13 - 0.97)** |
| Naproxen | NSAID | 978 | 6,352 | 0.5 | 1.5 | 0.4 | (0.13 - 1.22) |
| *A Streptococcus pneumoniae serotype X | vaccine | 385 | 6,945 | 1.4 | 2.6 | 0.42 | (0.13 - 1.35) |
| *B Streptococcus pneumoniae serotype X | vaccine | 609 | 6,721 | 1.5 | 2.5 | 0.46 | (0.18 - 1.19) |
| Flaxseed extract | supplement | 185 | 7,145 | 2.3 | 3.8 | 0.62 | (0.16 - 2.38) |
| Fluticasone propionate | corticosteroid | 895 | 6,435 | 0.9 | 1.4 | 0.63 | (0.24 - 1.69) |
| *C Streptococcus pneumoniae type X | vaccine | 505 | 6,825 | 1.9 | 2.8 | 0.65 | (0.26 - 1.66) |
| Sildenafil | PDE5 inhibitor | 180 | 7,150 | 2.8 | 4.2 | 0.68 | (0.19 - 2.37) |
| Dextromethorphan | antitussive | 899 | 6,431 | 0.7 | 1.1 | 0.68 | (0.22 - 2.06) |
| Amoxicillin | antibiotic | 1,706 | 5,624 | 0.8 | 1.1 | 0.68 | (0.30 - 1.54) |
| Clavulanate | antibiotic | 868 | 6,462 | 0.9 | 1.3 | 0.68 | (0.25 - 1.84) |
| Diclofenac sodium | NSAID | 336 | 6,994 | 1.8 | 2.5 | 0.72 | (0.24 - 2.22) |
| Thyrotropin | thyroid product | 468 | 6,862 | 1.1 | 1.4 | 0.74 | (0.21 - 2.58) |
| Morphine sulfate | opioid | 659 | 6,671 | 1.3 | 1.6 | 0.76 | (0.29 - 2.04) |
| Cetirizine | antihistamine | 1,384 | 5,946 | 0.9 | 1.2 | 0.78 | (0.33 - 1.82) |
| Fluticasone | corticosteroid | 2,155 | 5,175 | 1.1 | 1.4 | 0.79 | (0.41 - 1.54) |
| Omega-3 fatty acids | supplement | 430 | 6,900 | 2.1 | 2.6 | 0.79 | (0.30 - 2.11) |
| Tropicamide | opthalmic agent | 346 | 6,984 | 1.7 | 2 | 0.8 | (0.25 - 2.60) |
| Fexofenadine | antihistamine | 559 | 6,771 | 1 | 1.4 | 0.81 | (0.26 - 2.56) |
| Dexamethasone | corticosteroid | 881 | 6,449 | 1.1 | 1.5 | 0.82 | (0.33 - 2.06) |
| Vitamin B12 | supplement | 662 | 6,668 | 1.7 | 1.9 | 0.84 | (0.34 - 2.05) |
| Fish oils | supplement | 434 | 6,896 | 1.8 | 2.2 | 0.84 | (0.30 - 2.33) |
| Calcium carbonate | supplement | 307 | 7,023 | 1.6 | 2 | 0.84 | (0.23 - 3.01) |
| Vitamin D | supplement | 1,408 | 5,922 | 1.2 | 1.4 | 0.85 | (0.40 - 1.81) |
| Ketorolac | NSAID | 551 | 6,779 | 1.1 | 1.4 | 0.86 | (0.27 - 2.71) |
| Diclofenac | NSAID | 604 | 6,726 | 1.8 | 2.2 | 0.86 | (0.36 - 2.06) |
| Meloxicam | NSAID | 591 | 6,739 | 1.5 | 1.9 | 0.86 | (0.33 - 2.26) |
| Loratadine | antihistamine | 757 | 6,573 | 1 | 1.3 | 0.87 | (0.31 - 2.40) |
| Iron | supplement | 1,230 | 6,100 | 1.1 | 1.1 | 0.89 | (0.37 - 2.14) |
| Azelastine | antihistamine | 479 | 6,851 | 1.4 | 1.7 | 0.89 | (0.30 - 2.67) |
| Pseudoephedrine | decongestant | 953 | 6,377 | 0.7 | 1.1 | 0.91 | (0.31 - 2.65) |
| Methylprednisolone | corticosteroid | 869 | 6,461 | 1.3 | 1.6 | 0.92 | (0.38 - 2.21) |

| | | | | | | | |
|---|---|---|---|---|---|---|---|
| Cephapirin | antibiotic | 520 | 6,810 | 1.5 | 1.4 | 0.92 | (0.31 - 2.77) |
| Doxycycline | antibiotic | 690 | 6,640 | 1.4 | 1.5 | 0.93 | (0.36 - 2.40) |
| Ibuprofen | NSAID | 3,516 | 3,814 | 1 | 1.2 | 0.93 | (0.49 - 1.77) |
| Trazodone | antidepressant | 337 | 6,993 | 1.8 | 1.7 | 0.95 | (0.28 - 3.24) |
| Oxymetazoline | adrenergic agonist agent | 1,140 | 6,190 | 1.2 | 1.2 | 0.95 | (0.41 - 2.20) |
| Amlodipine | antihypertensive | 480 | 6,850 | 2.9 | 3.3 | 0.96 | (0.42 - 2.18) |
| Zolpidem | hypnotic | 240 | 7,090 | 2.1 | 2.1 | 0.97 | (0.25 - 3.68) |
| Tamsulosin | alpha 1 blocker | 214 | 7,116 | 4 | 4.1 | 0.97 | (0.33 - 2.83) |
| Folic acid | supplement | 416 | 6,914 | 1.7 | 1.5 | 0.97 | (0.31 - 3.08) |
| Varicella zoster virus glycoprotein E | vaccine | 221 | 7,109 | 2.6 | 3.1 | 0.97 | (0.27 - 3.47) |
| Ascorbic acid | supplement | 567 | 6,763 | 1.7 | 1.7 | 0.99 | (0.38 - 2.59) |

Other abbreviations and acronyms. *A Streptococcus pneumoniae serotype X: Streptococcus pneumoniae serotype (1, 19A, 3, 5, 6A, 7F) capsular antigen diphtheria CRM197 protein conjugate vaccine; *B Streptococcus pneumoniae serotype X: Streptococcus pneumoniae serotype (14, 18C, 19F, 23F, 4, 6B, 9V) capsular antigen diphtheria CRM197 protein conjugate vaccine; *C Streptococcus pneumoniae type X: Streptococcus pneumoniae type (1, 10A, 11A, 12F, 14, 15B, 17F, 18C, 19A, 19F, 2, 20, 22F, 23F, 3, 33F, 4, 5, 6B, 7F, 8, 9N, 9V) capsular polysaccharide antigen; NSAID: nonsteroidal anti-inflammatory drug; PDE5 inhibitor: phosphodiesterase type 5 inhibitor.

**eTable 10** Association between drug exposure and hospitalized-mild (exclusive severity).

**TCe/u**, total count of drug exposed/unexposed patients; **SRe/u**, severity rate of drug exposed/unexposed patients computed after overlap weighting with propensity score; **OR**, odds ratio; **CI**, confidence interval. The association results are arranged in the ascending order of OR values. The statistically significant results at the 0.05 level are emphasized in bold.

| Drug name | Drug class | TCe | TCu | SRe | SRu | OR | 95% CI |
|---|---|---|---|---|---|---|---|
| **Turmeric extract** | **supplement** | **150** | **7,474** | **3.3** | **10.4** | **0.32** | **(0.11 - 0.97)** |
| Ethinyl estradiol | estrogen | 748 | 6,876 | 0.8 | 2.3 | 0.4 | (0.14 - 1.17) |
| **Flaxseed extract** | **supplement** | **197** | **7,427** | **7.9** | **17.3** | **0.4** | **(0.19 - 0.84)** |
| ***C Streptococcus pneumoniae type X** | **vaccine** | **529** | **7,095** | **6.1** | **12.6** | **0.42** | **(0.25 - 0.69)** |
| Clotrimazole | antifungal | 158 | 7,466 | 3.1 | 6.6 | 0.44 | (0.13 - 1.43) |
| **Methylprednisolone acetate** | **corticosteroid** | **407** | **7,217** | **3.2** | **7.5** | **0.46** | **(0.22 - 0.94)** |
| Eicosapentaenoate | supplement | 229 | 7,395 | 5 | 9.6 | 0.5 | (0.23 - 1.12) |
| ***A Streptococcus pneumoniae serotype X** | **vaccine** | **407** | **7,217** | **6.6** | **11** | **0.52** | **(0.30 - 0.91)** |
| Glucosamine | supplement | 109 | 7,515 | 5.4 | 11.2 | 0.52 | (0.17 - 1.61) |
| Estradiol | estrogen derivative | 572 | 7,052 | 2.5 | 4.8 | 0.52 | (0.25 - 1.08) |
| Psyllium | antidiarrheal | 113 | 7,511 | 4.4 | 8.3 | 0.54 | (0.16 - 1.83) |
| **Omega-3 fatty acids** | **supplement** | **456** | **7,168** | **7.2** | **11.5** | **0.56** | **(0.33 - 0.95)** |
| Ubidecarenone | supplement | 130 | 7,494 | 6.8 | 12.1 | 0.56 | (0.23 - 1.41) |
| **Pseudoephedrine** | **decongestant** | **970** | **6,654** | **2.5** | **4.7** | **0.57** | **(0.33 - 0.99)** |
| Triamcinolone acetonide | corticosteroid | 154 | 7,470 | 5.1 | 8.3 | 0.57 | (0.21 - 1.56) |
| Biotin | supplement | 206 | 7,418 | 5.8 | 9.2 | 0.58 | (0.25 - 1.32) |
| Estrogens | estrogen | 314 | 7,310 | 3.7 | 6.6 | 0.58 | (0.26 - 1.29) |
| ***B Streptococcus pneumoniae serotype X** | **vaccine** | **647** | **6,977** | **7** | **10.4** | **0.59** | **(0.38 - 0.93)** |
| Clobetasol | corticosteroid | 151 | 7,473 | 5.2 | 7.7 | 0.59 | (0.22 - 1.62) |
| Azelastine | antihistamine | 495 | 7,129 | 4.5 | 7.4 | 0.61 | (0.33 - 1.10) |
| Betamethasone | corticosteroid | 244 | 7,380 | 4.9 | 7.8 | 0.62 | (0.28 - 1.36) |
| Oseltamivir | antiviral | 554 | 7,070 | 2.9 | 4.5 | 0.64 | (0.32 - 1.27) |
| Calcium citrate | calcium salt | 113 | 7,511 | 5.3 | 8.8 | 0.64 | (0.22 - 1.92) |
| Varicella zoster virus glycoprotein E | vaccine | 233 | 7,391 | 7.3 | 12.9 | 0.65 | (0.32 - 1.31) |
| **Ibuprofen** | **NSAID** | **3,608** | **4,016** | **3.7** | **5.6** | **0.65** | **(0.47 - 0.90)** |
| Estrogens, conjugated (USP) | estrogens | 118 | 7,506 | 5.9 | 9.4 | 0.65 | (0.22 - 1.90) |
| Nitrofurantoin, monohydrate | antibiotic | 207 | 7,417 | 3.4 | 5.1 | 0.65 | (0.22 - 1.94) |
| Cetirizine | antihistamine | 1,423 | 6,201 | 3.5 | 5.2 | 0.68 | (0.44 - 1.03) |
| **Fluticasone** | **corticosteroid** | **2,232** | **5,392** | **4.2** | **6.1** | **0.68** | **(0.48 - 0.95)** |
| Lutein | supplement | 157 | 7,467 | 10.7 | 14.7 | 0.71 | (0.34 - 1.48) |
| Lamotrigine | anticonvulsant | 157 | 7,467 | 3.2 | 4.4 | 0.72 | (0.21 - 2.47) |
| Diethylstilbestrol | estrogen | 182 | 7,442 | 9.7 | 13.2 | 0.72 | (0.35 - 1.47) |
| Testosterone | androgen | 293 | 7,331 | 5 | 7.1 | 0.72 | (0.34 - 1.52) |
| Fexofenadine | antihistamine | 579 | 7,045 | 4.4 | 6.1 | 0.73 | (0.41 - 1.30) |

| | | | | | | | |
|---|---|---|---|---|---|---|---|
| Naproxen | NSAID | 1,017 | 6,607 | 4.2 | 5.9 | 0.74 | (0.47 - 1.16) |
| Dextromethorphan | antitussive | 926 | 6,698 | 3.6 | 4.9 | 0.75 | (0.45 - 1.26) |
| Fluticasone propionate | corticosteroid | 932 | 6,692 | 4.7 | 5.9 | 0.76 | (0.48 - 1.20) |
| Tetanus toxoid vaccine, inactivated | vaccine | 1,218 | 6,406 | 4.6 | 5.9 | 0.76 | (0.50 - 1.14) |
| Acellular pertussis vaccine, inactivated | vaccine | 1,166 | 6,458 | 4.7 | 5.9 | 0.77 | (0.51 - 1.15) |
| Benzonatate | antitussive | 662 | 6,962 | 5.7 | 7.5 | 0.77 | (0.47 - 1.25) |
| Mometasone | corticosteroid | 214 | 7,410 | 5.1 | 7 | 0.77 | (0.32 - 1.83) |
| Vitamin B12 | supplement | 702 | 6,922 | 6.8 | 8.4 | 0.77 | (0.49 - 1.22) |
| Diphtheria toxoid vaccine, inactivated | vaccine | 1,216 | 6,408 | 4.7 | 5.9 | 0.77 | (0.52 - 1.16) |
| Naproxen sodium | NSAID | 134 | 7,490 | 5.9 | 7.4 | 0.78 | (0.28 - 2.19) |
| Haemophilus influenzae type b | vaccine | 371 | 7,253 | 4 | 5 | 0.78 | (0.37 - 1.66) |
| Oxymetazoline | adrenergic agonist agent | 1,179 | 6,445 | 4.4 | 5.3 | 0.79 | (0.51 - 1.21) |
| Loratadine | antihistamine | 785 | 6,839 | 4.5 | 5.7 | 0.79 | (0.48 - 1.31) |
| Varicella-zoster virus vaccine live (Oka-Merck) strain | vaccine | 158 | 7,466 | 5.7 | 6 | 0.79 | (0.29 - 2.15) |
| Metoprolol succinate | antihypertensive | 181 | 7,443 | 10.1 | 12.5 | 0.8 | (0.39 - 1.63) |
| Fish oils | supplement | 460 | 7,164 | 7.1 | 9.1 | 0.8 | (0.47 - 1.36) |
| *D Neisseria meningitidis serogroup X | vaccine | 171 | 7,453 | 2.9 | 3.5 | 0.82 | (0.23 - 2.99) |
| Fiber | supplement | 681 | 6,943 | 5 | 6.2 | 0.85 | (0.51 - 1.40) |
| Diclofenac sodium | NSAID | 362 | 7,262 | 8.3 | 10.3 | 0.85 | (0.49 - 1.48) |
| Docosahexaenoate | supplement | 176 | 7,448 | 5.7 | 6.6 | 0.85 | (0.34 - 2.13) |
| Sodium sulfate | laxative | 188 | 7,436 | 8.4 | 10.2 | 0.86 | (0.40 - 1.84) |
| Sildenafil | PDE5 inhibitor | 196 | 7,428 | 10.9 | 13.1 | 0.86 | (0.43 - 1.74) |
| Norethindrone | contraceptive | 438 | 7,186 | 1.9 | 2.5 | 0.86 | (0.31 - 2.41) |
| Tadalafil | PDE5 inhibitor | 128 | 7,496 | 10.4 | 13.2 | 0.87 | (0.38 - 2.01) |
| Cortisone | corticosteroid | 175 | 7,449 | 7.3 | 8.7 | 0.89 | (0.39 - 2.03) |
| Thioguanine | antineoplastic agent | 242 | 7,382 | 8.8 | 10 | 0.89 | (0.46 - 1.73) |
| Tretinoin | antineoplastic agent | 228 | 7,396 | 2.7 | 3.4 | 0.9 | (0.28 - 2.86) |
| Methylprednisolone | corticosteroid | 911 | 6,713 | 5.8 | 6.8 | 0.92 | (0.60 - 1.40) |
| Diclofenac | NSAID | 646 | 6,978 | 7.7 | 8.7 | 0.94 | (0.60 - 1.46) |
| Hydrochlorothiazide | antihypertensive | 767 | 6,857 | 9.7 | 11.1 | 0.96 | (0.65 - 1.41) |
| Ketoconazole | antifungal | 239 | 7,385 | 5.8 | 5.9 | 0.96 | (0.43 - 2.15) |
| Meloxicam | NSAID | 627 | 6,997 | 6.9 | 7.9 | 0.96 | (0.60 - 1.54) |
| Phenol | anesthetic | 349 | 7,275 | 4.3 | 4.4 | 0.96 | (0.44 - 2.10) |
| Hepatitis A vaccine (inactivated) strain HM175 | vaccine | 185 | 7,439 | 5.4 | 5.1 | 0.97 | (0.36 - 2.63) |
| Methocarbamol | skeletal muscle relaxant | 163 | 7,461 | 7.3 | 8.3 | 0.98 | (0.40 - 2.39) |
| Lufenuron | antiparasitic/antifungal | 213 | 7,411 | 7.9 | 7.6 | 0.99 | (0.47 - 2.10) |
| Ascorbic acid | supplement | 604 | 7,020 | 7.5 | 7.5 | 1 | (0.62 - 1.60) |
| Guaifenesin | expectorant | 1,260 | 6,364 | 6 | 5.9 | 1 | (0.68 - 1.47) |
| Amlodipine | antihypertensive | 530 | 7,094 | 11.3 | 12.2 | 1 | (0.65 - 1.54) |

Other abbreviations and acronyms. *A Streptococcus pneumoniae serotype X: Streptococcus pneumoniae serotype (1, 19A, 3, 5, 6A, 7F) capsular antigen diphtheria CRM197 protein conjugate vaccine; *B Streptococcus pneumoniae serotype X: Streptococcus pneumoniae serotype (14, 18C, 19F, 23F, 4, 6B, 9V) capsular antigen diphtheria CRM197 protein conjugate vaccine; *C Streptococcus pneumoniae type X: Streptococcus pneumoniae type (1, 10A, 11A, 12F, 14, 15B, 17F, 18C, 19A, 19F, 2, 20, 22F, 23F, 3, 33F, 4, 5, 6B, 7F, 8, 9N, 9V) capsular polysaccharide antigen; *D Neisseria meningitidis serogroup X: Neisseria meningitidis serogroup (C, W-135, Y) capsular polysaccharide diphtheria toxoid protein conjugate vaccine; NSAID: nonsteroidal anti-inflammatory drug; PDE5 inhibitor: phosphodiesterase type 5 inhibitor.
